# Prediction of future cognitive decline among cognitively unimpaired individuals using measures of soluble phosphorylated tau or tau tangle pathology

**DOI:** 10.1101/2024.06.12.24308824

**Authors:** Rik Ossenkoppele, Gemma Salvadó, Shorena Janelidze, Alexa Pichet Binette, Divya Bali, Linda Karlsson, Sebastian Palmqvist, Niklas Mattsson-Carlgren, Erik Stomrud, Joseph Therriault, Nesrine Rahmouni, Pedro Rosa-Neto, Emma M. Coomans, Elsmarieke van de Giessen, Wiesje M. van der Flier, Charlotte E. Teunissen, Erin M. Jonaitis, Sterling C. Johnson, PREVENT-AD Research Group, Sylvia Villeneuve, Tammie L.S. Benzinger, Suzanne E. Schindler, Randall J. Bateman, James D. Doecke, Vincent Doré, Azadeh Feizpour, Colin L. Masters, Christopher Rowe, Heather J. Wiste, Ronald C. Petersen, Clifford R. Jack, Oskar Hansson

## Abstract

Plasma p-tau217 and Tau-PET are strong prognostic biomarkers in Alzheimer’s disease (AD), but their relative performance in predicting future cognitive decline among cognitively unimpaired (CU) individuals is unclear. In this head-to-head comparison study including 9 cohorts and 1534 individuals, we found that plasma p-tau217 and medial temporal lobe Tau-PET signal showed similar associations with cognitive decline on a global cognitive composite test (R^2^_PET_=0.32 vs R^2^_PLASMA_=0.32, p_difference_=0.812) and with progression to mild cognitive impairment (Hazard ratio[HR]_PET_=1.56[1.43-1.70] vs HR_PLASMA_=1.63[1.50-1.77], p_difference_=0.627). Combined plasma and PET models were superior to the single biomarker models (R^2^=0.36, p<0.01). Furthermore, sequential selection using plasma p-tau217 and then Tau-PET reduced the number of participants required for a clinical trial by 94%, compared to a 75% reduction when using plasma p-tau217 alone. We conclude that plasma p-tau217 and Tau-PET showed similar performance for predicting future cognitive decline in CU individuals, and their sequential use (i.e., plasma p-tau217 followed by Tau-PET in a subset with high plasma p-tau217) is useful for screening in clinical trials in preclinical AD.

---

In recent years, there has been a substantial increase in the availability of biomarkers that reflect the core Alzheimer’s disease (AD) neuropathological hallmarks, specifically Amyloid-β (Ab) plaques and tau neurofibrillary tangles.^1^ Tau-PET has shown excellent diagnostic accuracy and strongly associations with both concurrent and longitudinal cognitive decline, outperforming established AD biomarkers like Amyloid-PET and structural MRI across the clinical continuum of AD.^1–7^ However, Tau-PET is expensive and labor intensive, has inadequate availability and its sensitivity to detect the earliest stages of tau aggregation is limited. This is particularly troublesome in individuals with preclinical AD who harbor AD pathology but have not (yet) developed symptoms.^8^ The recent advent of blood-based biomarkers of AD pathology potentially offers a low cost, non-invasive and scalable alternative.^9^ Within the swiftly evolving realm of plasma biomarkers, plasma p-tau217 (phosphorylated-tau at threonine 217) has demonstrated excellent performance in detecting AD pathology, distinguishing between AD and non-AD neurodegenerative disorders, and predicting future clinical progression.^10–16^ However, in contrast with Tau-PET, plasma p-tau217 provides no regional information on AD pathology, its continuous values are less representative of the full dynamic range of tau pathology and its signal represents a mix of tau and Aβ pathology and is therefore a less tau-specific biomarker.^17–19^ In cognitively impaired individuals, Tau-PET has shown superior performance to plasma p-tau217 in predicting future cognitive decline.^20,21^ In cognitively unimpaired (CU) individuals, however, it is yet unclear whether there is a meaningful difference in prognostic utility between Tau-PET and plasma p-tau217 biomarkers.^22^ Determining which of the two biomarkers is the strongest predictor of future cognitive deterioration in initially CU individuals is of utmost importance as clinical trials are increasingly recruiting participants with preclinical AD to enable early intervention. This information would become even more crucial if treatments such as lecanemab^23^ and donanemab^24^ are found to be effective in preclinical AD, as this would require large-scale screening of CU populations for AD pathology. Therefore, we performed a large-scale head-to-head comparison study between Tau-PET (from which signal was extracted from reporter regions covering the medial temporal lobe [Tau-PET_MTL_] and the temporal neocortex [Tau-PET_NEO_]^5^) vs plasma p-tau217. We assessed their associations with longitudinal cognitive decline and diagnostic progression to mild cognitive impairment (MCI). Additionally, we examined whether and how plasma p-tau217 and Tau-PET can be combined to further increase their prognostic accuracy and to optimize recruitment strategies for clinical trials.

## RESULTS

### Participants

We included 1534 CU participants from nine cohorts with Tau-PET and plasma p-tau217 data available at baseline, of whom 413 (26.7%) were Aβ−positive on PET (see **Table-1** and **Extended-Data Table-1** for data by cohort). The mean±standard deviation age of the participants was 68.4±10.0 years, 53.8% were females and the follow-up duration was 3.8±1.8 years. The associations between Tau-PET and plasma p-tau217 levels were moderate (plasma p-tau217 vs Tau-PET_MTL_, ρ[95%CI] = 0.42 [0.38-0.46], p<0.001; plasma p-tau217 vs Tau-PET_NEO_, ρ=0.34 [0.30-0.39], p<0.001, **Extended-Data Figure-1**).

**Table 1.**
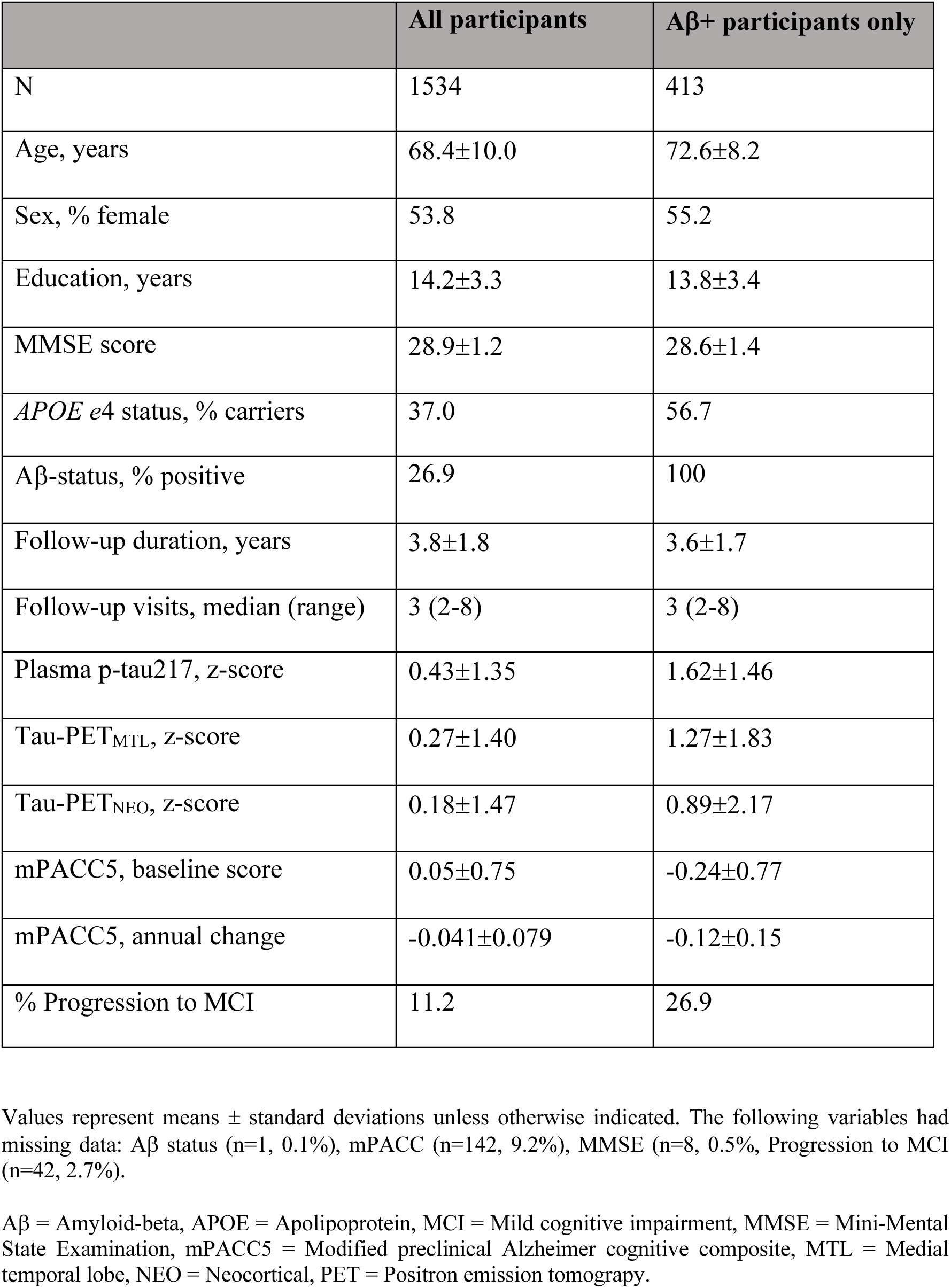
Participant characteristics.

### Prediction of future decline in cognitive function

First, we examined across all participants whether the tau biomarkers individually and combined were associated with cognitive decline over time on the modified preclinical Alzheimer cognitive composite (mPACC5). We selected the mPACC5 because it is a sensitive measure that can reliably detect longitudinal changes over time in CU populations, and is therefore often used as an outcome measure in research studies and clinical trials focusing on preclinical AD.^25,26^ mPACC5 slopes were generated using linear mixed effects models and then used as dependent variable in linear regression models adjusting for age, sex, years of education, *APOE* ε4 carriership and cohort. The analysis showed that plasma p-tau217 concentrations (R^2^[95%CI]= 0.32 [0.27-0.35], corrected Akaike Information Criterion [AICc]=-3766), Tau-PET uptake in the medial temporal lobe (Tau-PET_MTL_, R^2^= 0.32 [0.27-0.36], AICc= -3773 and Tau-PET uptake in the temporal neocortex (Tau-PET_NEO_, R^2^= 0.31 [0.25-0.35], AICc= -3751) were all better predictors of longitudinal cognitive decline than basic models that included age, sex, education and cohort with *APOE* ε4 status (R^2^= 0.24 [0.20-0.27], AICc= -3617) or without *APOE* ε4 status (R^2^= 0.23 [0.19-0.26], AICc= -3603) (**Figure-1a-b**, **Extended-Data Table-2/3**). There were no significant differences between single biomarker models, i.e. between plasma p-tau217 and Tau-PET_MTL_ (p=0.812), between plasma p-tau217 and Tau-PET_NEO_ (p=0.699), and between Tau-PET_MTL_ and Tau-PET_NEO_ (p=0.404). Combined biomarker models, i.e., plasma p-tau217 and Tau-PET_MTL_ (R^2^= 0.36 [0.30-0.40], AICc= -3849) and plasma p-tau217 and Tau-PET_NEO_ (R^2^= 0.35 [0.29-0.40], AICc= -3841) were more strongly associated with longitudinal mPACC5 decline than the single biomarker models (all p<0.01). The relative contribution of each biomarker in the combined models further indicated their complementary value, as e.g., in the combined plasma p-tau217 and Tau-PET_MTL_ model, of all explained mPACC5 variance 15% was explained by plasma p-tau217 alone, 17% by Tau-PET_MTL_ alone, 24% was shared variance and the remaining 43% was explained by covariates (**Figure-1c**, **Extended-Data Table-4**). The results were overall consistent across cohorts (**Extended-Data Figure-2/3 and Extended-Data Table-5**).

**Figure 1.**
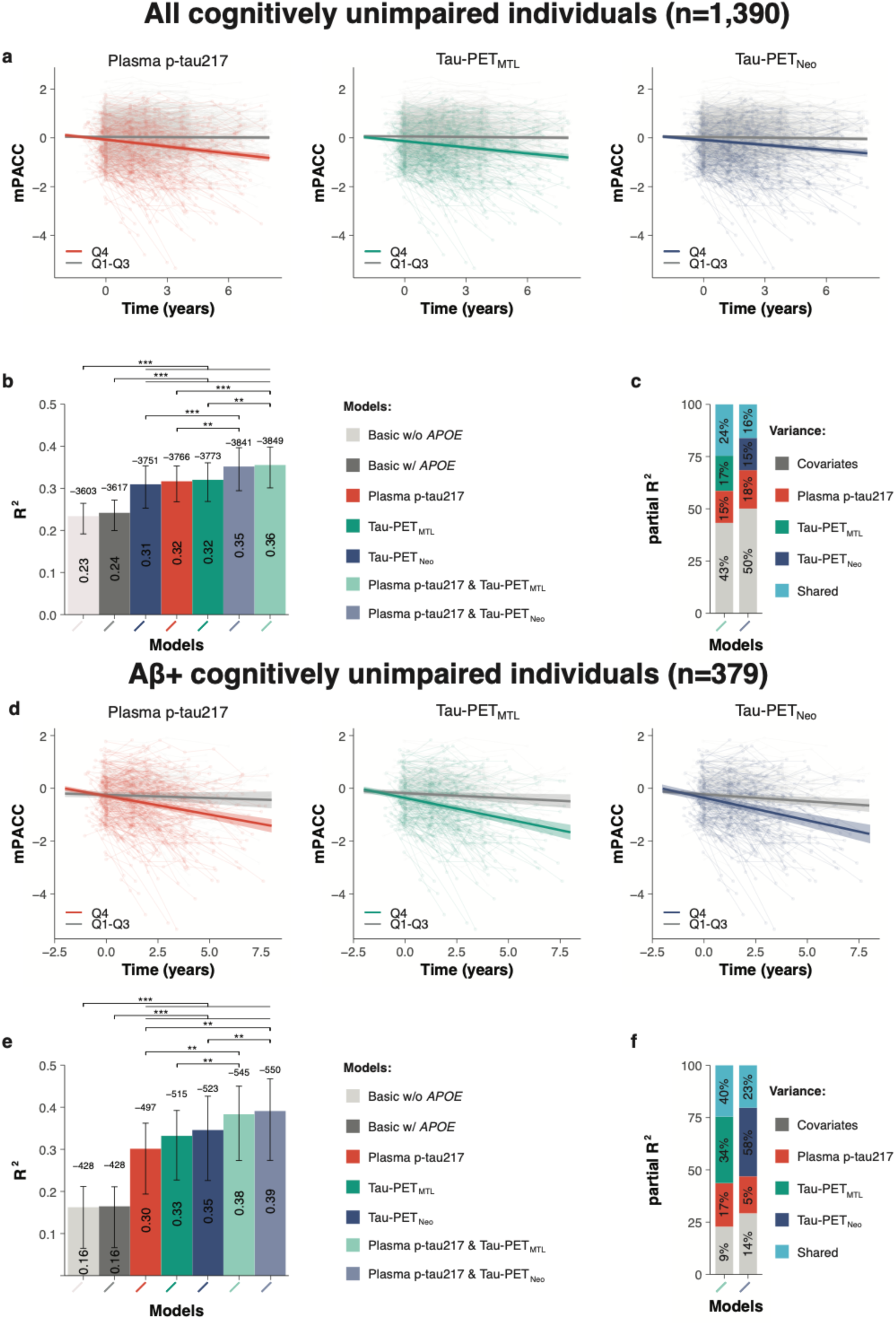
Plasma p-tau217 and Tau-PET prediction of future cognitive decline. **a**, **d,** Scatterplots showing the association between cognitive change over time on the mPACC5 and the tau biomarkers (Quartile 1-3 vs Quartile 4) across all participants (**a**) and Aβ+ participants (**d**) only. The shadow area indicated the 95% confidence interval. **b**, **e,** Explained variance (R^2^, inside the bar plot) and model fit (corrected Akaike criterion, outside the bar plot) for various models predicting longitudinal change on the mPACC5 across all participants (**b**) and Aβ+ (**d**) participants only. Errorbars represent the 95% CI. **c, f,** Partial explained variance (R^2^) for combined biofluid and neuroimaging models predicting longitudinal change on the mPACC5 across all participants (**c**) and Aβ+ (**f**) participants only. **p<0.01, ***p<0.001.

Next, we repeated the same set of analyses with mPACC5 as the outcome measure in Amyloid-PET positive CU individuals only (n=379). This analysis yielded a largely similar pattern compared to analyses in the entire sample (**Figure-1d-e, Extended-Data Table-2/3**). The single biomarker models including plasma p-tau217 (R^2^= 0.30 [0.19-0.36], AICc= -497), Tau-PET_MTL_ (R^2^= 0.33 [0.22-0.40], AICc= -515) or Tau-PET_NEO_ (R^2^= 0.35 [0.22-0.43], AICc= -523) outperformed the basic models with and without *APOE* ε4 status (R^2^=0.16[0.07-0.21], AICc= -428 for both). Further, there were no differences between the three single biomarker models (plasma p-tau217 vs Tau-PET_MTL_, p=0.344; plasma p-tau217 vs Tau-PET_NEO_, p=0.287; Tau-PET_MTL_ vs Tau-PET_NEO_, p=0.693). Also, combined biomarker models, i.e., plasma p-tau217 and Tau-PET_MTL_ (R^2^= 0.38 [0.27-0.45], AICc= -546) and plasma p-tau217 and Tau-PET_NEO_ (R^2^= 0.39 [0.27-0.47], AICc= -550), were more strongly associated with longitudinal mPACC5 decline than their respective single biomarker models (both p<0.01). Among Aβ-positive CU individuals, the relative contribution of the Tau-PET measures in the combined model was substantially greater than in the entire study population, i.e., Tau-PET_MTL_ 34% vs 17% and Tau-PET_NEO_ 58% vs 15%, which was not the case for plasma p-tau217 (15% vs 17%, **Figure-1c-f**, **Extended-Data Table-4**).

### Prediction of clinical progression to Mild Cognitive Impairment (MCI)

Next, we examined across all participants whether the tau biomarkers individually and combined were associated with the rate of clinical progression to MCI using Cox proportional hazard models, adjusting for age, sex, years of education, cohort and *APOE*ε4 carriership. The analysis revealed that higher baseline plasma p-tau217 (Hazard ratio[HR]= 1.57 [1.44-1.71], AICc= 2099), Tau-PET_MTL_ (HR= 1.63 [1.50-1.77], AICc= 2077) and Tau-PET_NEO_ (HR= 1.42 [1.33-1.51], AICc= 2111) levels were all associated with an increased risk for future progression to MCI (all p<0.001, **Figure-2a-b**, **Extended-Data Table-6/7**). The individual fluid vs neuroimaging tau biomarker models did not differ from each other (plasma p-tau217 vs Tau-PET_MTL_, p=0.340; plasma p-tau217 vs Tau-PET_NEO_, p=0.571), while the performance of Tau-PET_MTL_ was slightly better than that of Tau-PET_NEO_ (p=0.046). The fit was improved for the plasma p-tau217 model when adding Tau-PET_MTL_ (ΔAICc= -52, p=0.005) or Tau-PET_NEO_ (ΔAICc= -30, p=0.018, **Extended-Data Table-6/8**). Likewise, the model fit also improved when adding plasma p-tau217 to Tau-PET_MTL_ (ΔAICc= -30, p=0.007) and to Tau-PET_NEO_ (ΔAICc= - 42, p=0.001). The results were largely consistent across cohorts (**Extended-Data Figure-4/5**).

**Figure 2.**
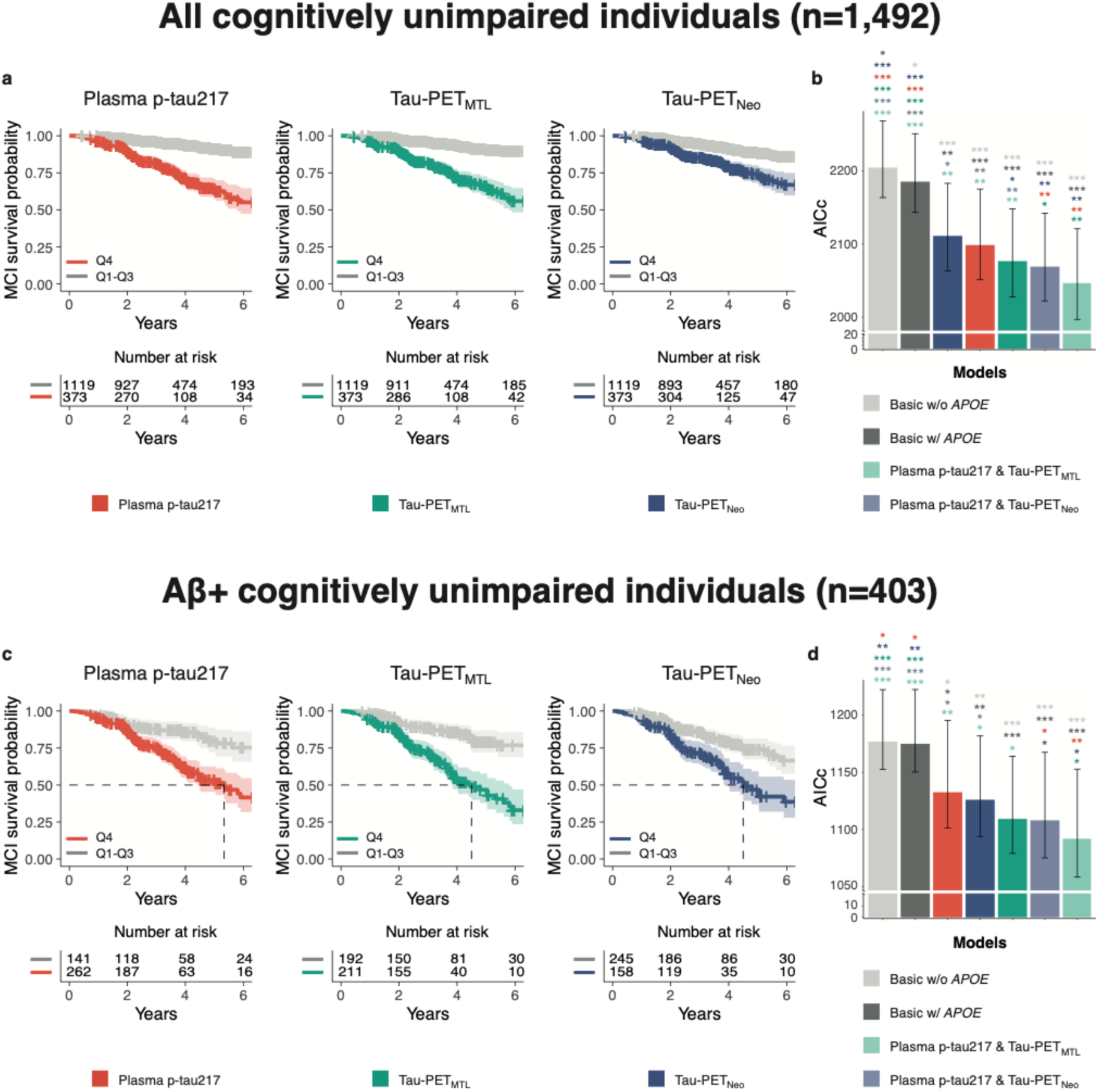
Plasma p-tau217 and Tau-PET prediction of progression to mild cognitive impairment. **a**, **c,** Survival curves for progression to mild cognitive impairment (Quartile 1-3 vs Quartile 4) across all participants (**a**) and Aβ+ (**c**) participants only, including a table of total number of participants available at each time point. The dashed line in **c** indicates the time point at which 50% of a group had progressed to MCI, and the shadow area indicated the 95% confidence interval. **b**, **d,** Model fit (corrected Akaike criterion) for various models predicting future clinical progression to mild cognitive impairment across all participants (**b**) and Aβ+ (**d**) participants only. Errorbars represent the 95% CI. * p<0.05, **p<0.01, ***p<0.001.

When repeating the same set of analyses in Aβ-positive CU individuals only (n=403), we found a largely similar pattern (**Figure-2c-d, Extended-Data Table-6/7**). Baseline plasma p-tau217 (HR= 1.56 [1.37-1.77], AICc= 1133), Tau-PET_MTL_ (HR= 1.54 [1.39-1.70], AICc=1109) and Tau-PET_NEO_ (HR= 1.34 [1.25-1.43], AICc= 1126) levels were all associated with an increased risk for future progression to MCI (all p<0.001). The individual Tau biomarker models did not differ from each other (plasma p-tau217 vs Tau-PET_MTL_, p=0.270; plasma p-tau217 vs Tau-PET_NEO_, p=0.754; Tau-PET_MTL_ vs Tau-PET_NEO_, p=0.244). Adding Tau-PET to plasma p-tau217 models improved the model fit when adding Tau-PET_MTL_ (ΔAICc= -41, p=0.002) and Tau-PET_NEO_ (ΔAICc= -25, p=0.03, **Extended-Data Table-6/8**). The model fit also slightly improved when adding plasma p-tau217 to either Tau-PET_MTL_ (ΔAICc= -17, p=0.043) or Tau-PET_NEO_ (ΔAICc= -18, p=0.049).

### A two-step approach to reduce the sample size in clinical trials

Then we tested whether and how a two-step sequential approach (i.e., plasma p-tau217 followed by Tau-PET) could reduce the number of participants needed for a preclinical AD trial using longitudinal changes in cognitive function as primary outcome. Significant sample size reductions can already be achieved in the first step, i.e., using only plasma p-tau217. When using the mPACC5 as outcome measure, assuming 80% power and α=0.05 in a 4-year clinical trial with annual repeated testing, selecting participants with plasma p-tau217 levels in Quartiles 2-4 (i.e., Q2-4, excluding the lowest 25% of plasma p-tau217) would result in a 32[14-41]% reduction of the number of required participants compared to including the entire study population (**Figure-3**). Selecting participants in plasma p-tau217 Q3-4 and Q4 further reduced the required sample size by 63[48-69]% and 81[72-86]%, respectively. Using clinical progression to MCI as an outcome measure yielded similar results, i.e., plasma p-tau217 Q2-4: 29[22-36]%, Q3-4: 56[48-63]% and Q4: 82[76-87]%, **Figure-4**. In the second step, Tau-PET measures further reduced the required sample size. For example, in the population with plasma p-tau217 concentrations in Quartiles 3-4, selecting participants with Tau-PET in Quartile 4 would further reduce the sample size from 63[48-69]% (plasma p-tau217) to 89[82-91]% (Tau-PET_MTL_, **Figure-3**) or to 85[77-89]% (Tau-PET_NEO_, **Extended Figure-6**) when using the mPACC5 as the outcome measure. Another example, in the population with plasma p-tau217 concentrations in Quartile 4, selecting participants with Tau-PET in Quartiles 3-4 would further reduce the sample size 82[76-87]% (plasma p-tau217) to 88[82-94]% (Tau-PET_MTL_, **Figure-4**) or to 94[91-97]% (Tau-PET_NEO_, **Extended Figure-7**) when using clinical progression to MCI as the outcome measure. The estimated sample size reductions for all plasma p-tau217 and Tau-PET quartile combinations are presented in **Extended Table-9**. Repeating the same set of analyses but now restricted to Aβ-positive CU individuals showed that similarly large sample size reductions can be achieved in this population (**Extended Figure-8**).

**Figure 3.**
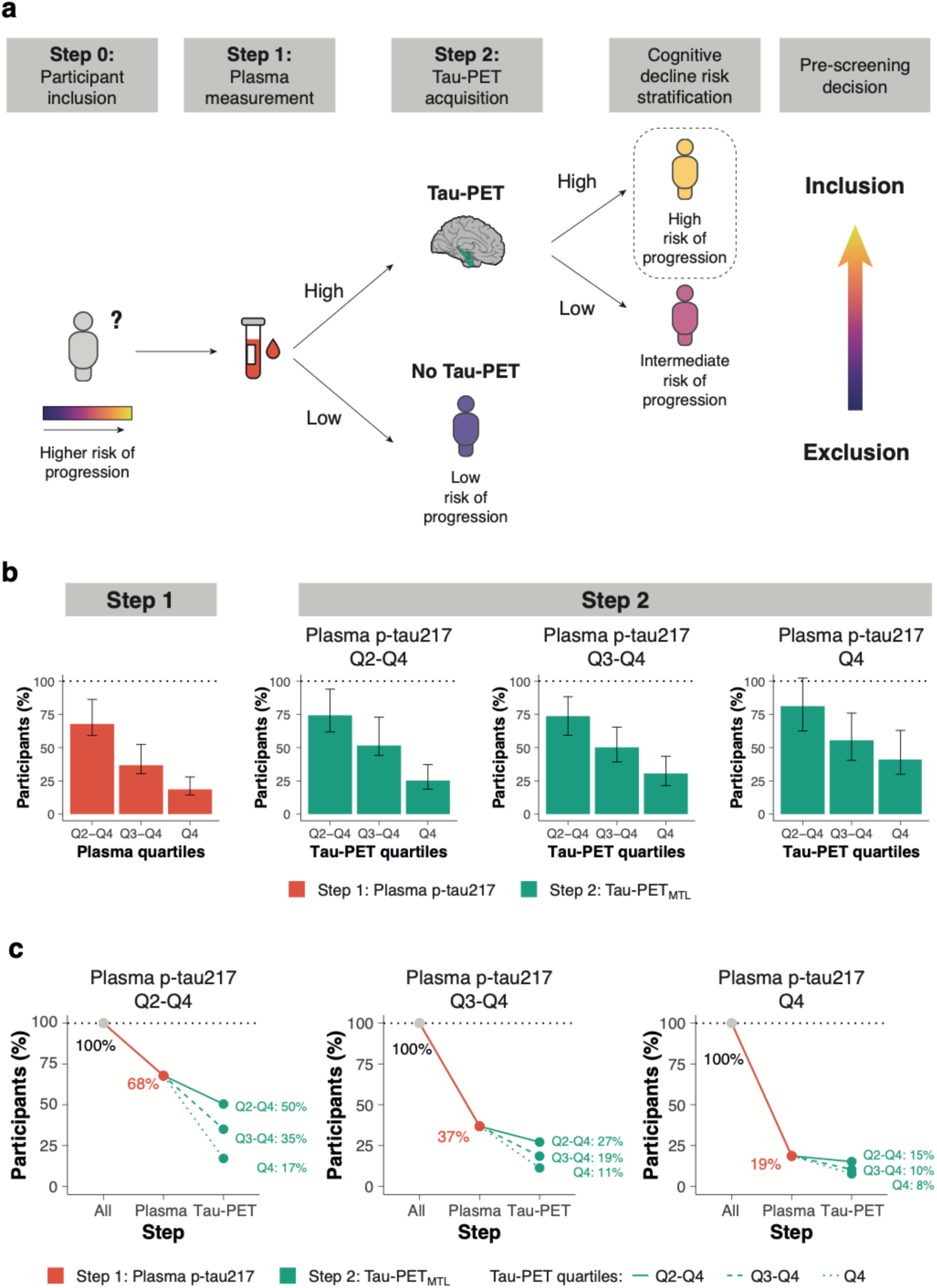
A two-step recruitment approach for clinical trials in preclinical Alzheimer’s disease using the mPACC as outcome measure. **a**, conceptual framework of a sequential two-step recruitment strategy of a clinical trial in preclinical Alzheimer’s disease using a cognitive endpoint. **b**, the obtained sample size reduction using sample selection based on different percentiles (75th, 50th and 25th) of baseline plasma p-tau217 levels in step 1 followed by the selection based on the same percentiles (75th, 50th and 25th) of the Tau-PET_MTL_ measurement in step 2 with mPACC5 as the primary endpoint. Note that 100% in step 2 refers to the participants selected by plasma p-tau217 in step 1. Errorbars represent the 95% CI. **c** shows the calculated sample size reductions for various plasma p-tau217 and Tau-PET_MTL_ quartile combinations

**Figure 4.**
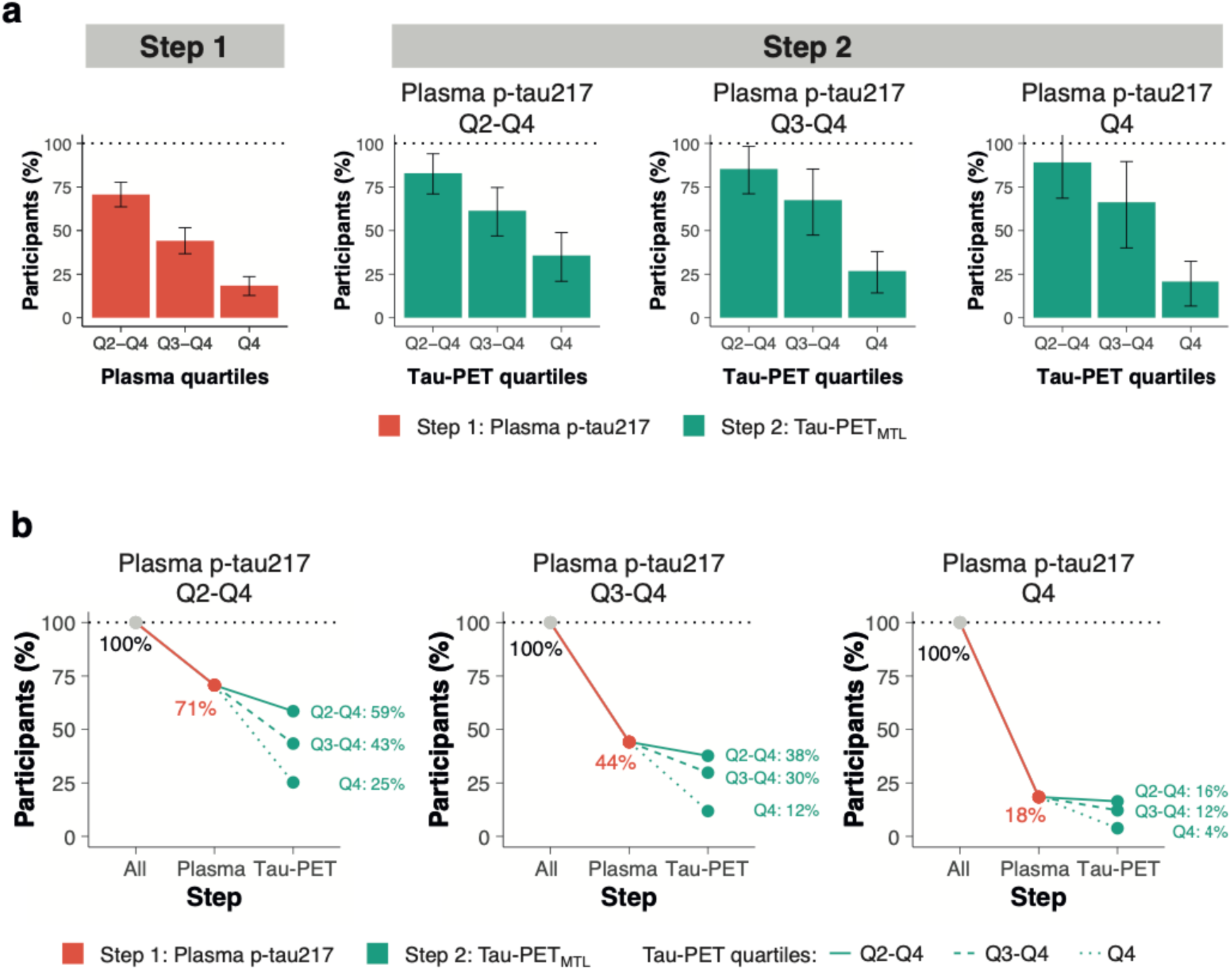
Clinical trial sample size reductions through a two-step recruitment strategy when using clinical progression to mild cognitive impairment as an outcome measure. **a**, the obtained sample size reduction using sample selection based on different percentiles (75th, 50th and 25th) of baseline plasma p-tau217 levels in step 1 followed by the selection based on the same percentiles (75th, 50th and 25th) of the Tau-PET_MTL_ measurement in step 2 with clinical progression to mild cognitive impairment as the primary endpoint. Note that 100% in step 2 refers to the participants selected by plasma p-tau217 in step 1. Errorbars represent the 95% CI. **b** shows the calculated sample size reductions for various plasma p-tau217 and Tau-PET_MTL_ quartile combinations.

### Characterization of different combined plasma p-tau217 and Tau-PET groups

Finally, we aimed to characterize the groups resulting from combining different plasma p-tau217 and Tau-PET_MTL_ quartiles in terms of Aβ-positivity, rates of mPACC5 decline and progression to MCI, as well as the number of participants that would be included based on each combination relative to the full dataset. For this purpose, we created four different groups based on increasingly restrictive tau biomarker combinations: A) a “liberal” group consisting of participants with plasma p-tau217 levels in Q2-Q4 and then Tau-PET_MTL_ uptake in Q2-Q4 of those selected by plasma, B) a “moderate” group consisting of plasma p-tau217 levels in Q3-Q4 and then Tau-PET_MTL_ uptake in Q3-Q4, C) a “plasma p-tau217 Q4 only” group consisting of individuals with plasma p-tau217 levels in Q4 regardless of Tau-PET, and D) a “conservative” group consisting of plasma p-tau217 levels in Q4 and then Tau-PET_MTL_ uptake in Q4. **Figure-5a-c** indicates a progressively worse outcome for individuals from approach A to D, with an increasing proportion of Aβ-positive individuals from the liberal (A) to the conservative (D) threshold approach (from 39.8% to 95.6%), more rapid decline on the mPACC5 (from standardized-β= -0.06±0.09 to -0.15±0.12) and a greater proportion of CU individuals who progressed to MCI (from 17.0% to 40.1%). Furthermore, the proportion of participants that would be included in a hypothesized clinical trial with the mPACC5 as an outcome measure decreased from 56.2% (of the entire population) when using the liberal threshold approach to only 6.2% when using the conservative threshold approach (**Figure-5d**). Notably, there were no group differences between the plasma p-tau217 Q4-only approach vs the moderate combined threshold approach for mPACC5 decline (standardized-β= -0.09±0.10 vs -0.09±0.10), proportion with progression to MCI (26.0% vs 26.5%) and the proportion of participants selected (25.0% vs 25.0%), and only a modest difference in Aβ-positivity (i.e., 61.4% vs 71.7%). The same set of analyses using Tau-PET_NEO_ instead of Tau-PET_MTL_ yielded similar results (**Extended Figure-9**). Detailed group characterizations for all the other possible combinations are presented in **Extended Table-10** (i.e., Aβ-positivity, mPACC5 decline and clinical progression) and **Extended Table-11** (i.e., age, sex, *APOE* status and years of education).

**Figure 5.**
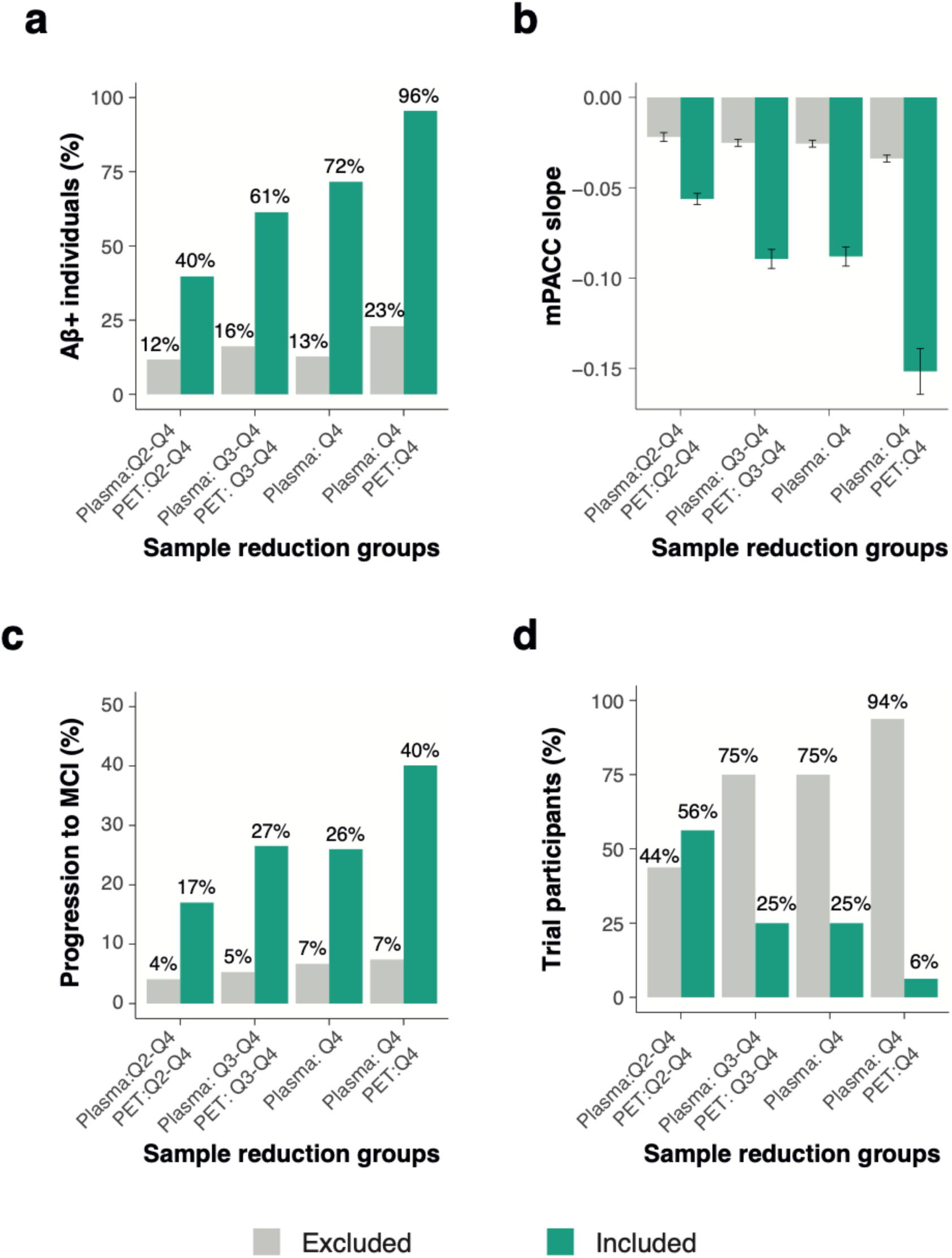
Characterization of different plasma p-tau217/Tau-PET_MTL_ groups on relevant trial measures. This figure shows how different group compositions based on their baseline plasma p-tau217 and Tau-PET_MTL_ levels are related to various relevant trial metrics, including the proportion of Aβ+ individuals (**a**), annual mPACC5 slope (**b**), proportion of initially cognitively unimpaired individuals that progress to mild cognitive impairment during a 4-year trial (**c**), and the proportion of individuals from the entire population that fall within the group definitions described on the x-axis (**d**). Errorbars in **b** represent the 95% CI.

We also calculated the projected costs that could be saved in a hypothetical trial with mPACC5 or MCI progression rates as an outcome measure using the same four groups as above. When assuming a 1:15 ratio (i.e., cost of 1 Tau-PET scan equals 15 plasma p-tau217 assessments) and using Tau-PET_MTL_, both the plasma p-tau217 Q4 only (C) group (96.9%) and the conservative combined (D) group (96.5% cost reduction) yielded substantially higher cost reducations compared to the moderate (B) group (85.4%) and especially the liberal (A) group (53.9%) when using mPACC as an endpoint (**Extended Figure-10a**, Tau-PET_NEO_ results are presented in **Extended Figure-10b**). Using the same ratio (1:15) but now using clinical progression as the endpoint, the plasma p-tau217 Q4 only (C) group yielded the highest cost reduction (88.2%), followed by the conservative combined (D) group (70.4%), the moderate (B) group (53.0%) the liberal (A) group (25.9%, **Extended Figure-11a**, Tau-PET_NEO_ results are presented in **Extended Figure-11b**).

## DISCUSSION

In this multicohort study, we investigated whether Tau-PET or plasma p-tau217 is more strongly associated with future cognitive decline among 1534 CU individuals and whether they would provide complementary information in screening approaches for clinical trials. This is a timely research question as both tau biomarkers are frequently incorporated into clinical trials, and often in combination as they reflect different aspects of tau pathophysiology. According to the draft revised AD criteria by the Alzheimer’s Association Workgroup, plasma p-tau217 is a Core 1 biomarker (T_1_) of phosphorylated and secreted AD tau, while Tau-PET is a Core 2 biomarker (T_2_) of AD Tau proteinopathy.^27^ In this study, we observed comparably strong associations between plasma p-tau217 and Tau-PET with cognitive decline on a sensitive global cognitive composite test (i.e., mPACC5) and with clinical progression to MCI in CU individuals. Importantly, models including both plasma p-tau217 and Tau-PET consistently outperformed single biomarker models, suggesting that plasma p-tau217 and Tau-PET provide complementary information. Simulations showed that a two-step approach (i.e., plasma p-tau217 first, followed by Tau-PET in individuals with high plasma p-tau217 only) could substantially reduce the number and cost of recruiting participants for a preclinical AD clinical trial with mPACC5 or progression to MCI as the primary endpoint. We conclude that plasma p-tau217 and Tau-PET showed similar associations with future cognitive decline in a CU population, that both tau biomarkers provide complementary information, and that their sequential use (i.e., plasma p-tau217 followed by Tau-PET in a subset with high plasma p-tau217) is useful for screening in clinical trials in preclinical AD.

A main finding of this study is that there were no statistical differences between plasma p-tau217 concentrations and Tau-PET uptake in their associations with future cognitive changes in a CU population. This is in contrast with cognitively impaired populations, where Tau-PET generally outperforms plasma p-tau217 in terms of prognostic accuracy.^20,21^ This discrepancy between disease stages might be explained by the differences in underlying pathophysiology and subsequent temporal dynamics of the two tau biomarkers. Plasma p-tau217 measures the hyperphosphorylated tau protein in soluble forms and has been shown to change very early in the disease process.^28^ Post-mortem studies indicated that ante-mortem plasma p-tau217 levels are associated with the density of both neurofibrillary tau tangle and Aβ plaque pathology in the brain.^18,29^ It is conceivable that this mix of Aβ- and tau-related signals reflected by plasma p-tau217 levels contributed to its non-inferiority versus Tau-PET for predicting cognitive change over time in this early population. For instance, Aβ toxicity might impact synaptic function or neuroinflammation, which could subsequently influence cognitive performance.^30^ In contrast, Tau-PET signal largely represents the presence of aggregated paired helical filaments of the tau protein forming insoluble neurofibrillary tangles.^31^ *In vivo* studies have shown that positive Tau-PET scans are relatively rare in CU individuals (i.e., ∼5-10% among Aβ-positive CU individuals in a temporal meta-ROI)^3,4^ and post-mortem studies have indicated that a positive [^18^F]Flortaucipir Tau-PET scan (quantitatively or by visual read) represents tau pathology in Braak stages IV and above.^32,33^ Altogether, this indicates that substantial changes on a Tau-PET scan occur in rather advanced clinical and biological stages of AD, which may explain why Tau-PET in the current mix of Aβ-positive and Aβ-negative CU individuals did not outperform plasma p-tau217 in predicting future cognitive decline. We also observed no marked differences between Tau-PET_MTL_ and Tau-PET_NEO_, which may appear contradictory with our previous study where the Tau-PET_NEO_-positive group exhibited a considerably worse prognosis.^34^ This discrepancy can be explained by the different group definitions used in the current study (quartile-based) compared to the previous (binary cut-offs). In the previous study, the Tau-PET_NEO_ positive group comprised a relatively small proportion of the overall CU population (∼4%). As a result, even in the highest quartile (Q4) of the current study, most individuals are Tau-PET negative. This attenuates the overall association between Tau-PET and cognitive decline, and dilutes the association even further for the Tau-PET_NEO_ group.

Since there were no clear distinctions between the two tau biomarkers in individual models, plasma p-tau217 should be prioritized over Tau-PET as a standalone screening tool in CU populations due to its major practical advantages. However, simultaneous modelling of plasma p-tau217 and Tau-PET consistently led to a more accurate forecast of subsequent cognitive decline compared to single biomarker models. We therefore tested a two-step sequential approach, which involved selecting participants based on plasma p-tau217 as the first step and then performing Tau-PET as the second step. This approach decreased the cost of selecting appropriate participants for clinical trials by drastically reducing the number of required Tau-PET scans and participants for screening. This was mainly achieved by selecting from the entire CU study population the subset of individuals that are at highest-risk for short-term cognitive decline due to their elevated levels of both baseline plasma p-tau217 and Tau-PET. In such a workflow (see **Figure-3a**), the participants’ eligibility in terms of inclusion (e.g., age, cognitive status) and exclusion (e.g., the absence of major neurological or psychiatric disorders) criteria would be assessed first. Next, participants would be further triaged based on their plasma p-tau217 levels, where “low/negative” participants would not undergo a Tau-PET scan as their risk for future cognitive decline would be low, while “high/positive” participants would undergo a Tau-PET scan to further refine their risk profile.^35^ The degree of Tau-PET uptake can then be utilized to assign CU individuals on a continuum ranging from intermediate (i.e., “high/positive” plasma p-tau217, “low/negative” Tau-PET) to high (i.e., “high/positive” results on both plasma p-tau217 and Tau-PET) risk for future progression. The final pre-screening decision will be based on factors such as the acceptable rate of screening failures, trial duration, expected effect size of the drug or intervention and whether the trial is a primary or secondary intervention strategy. The framework depicted in **Figure-3a** is primarily conceptual, and we recognize that numerous crucial decisions need to be made during its implementation. This includes, among other factors, determining the criteria for “high/positive” vs “low/negative” for both plasma p-tau217 and Tau-PET, deciding whether to use p-tau217 alone or in combination with other plasma biomarkers (e.g., Aβ42/40), identifying the target region-of-interest for Tau-PET quantification and selecting a quantitative threshold and/or visual read metric for Tau-PET.^36,37^

The main strength of this study is the multicenter approach that yielded a sufficient sample size for a robust head-to-head comparison between Tau-PET and plasma p-tau217 as well as a thorough assessment of their potential complementary value. The main limitations of the study are related to the inherent challenges of a multi-center study that was not co-designed from the start. We aimed to optimize pooling of data across cohorts by standardizing biomarkers values using CU Aβ-negative participants. We acknowledge, however, that there may still be residual heterogeneity caused by use of different Amyloid-PET and Tau-PET tracers and different p-tau217 assays, as well as the use of different neuropsychological tests to generate mPACC5 scores. To mitigate this, we present both the pooled (main report) and cohort-specific (supplement) results. Moreover, we did not have plasma p-tau217 cut-offs available from all cohorts and instead used either continuous (z-scores) or categorical (quartiles) data for the analyses. Additionally, Tau-PET may have been at a slight disadvantage compared to plasma p-tau217, as Tau-PET data were more often analyzed locally, whereas plasma p-tau217 was predominantly analyzed centrally at Lund University. Future studies using predefined cut-offs for both plasma p-tau217 and Tau-PET, preferentially in a more diverse population in terms of ethnicity, socioeconomic status and medical comorbidities, are of importance to establish the generalizability of our findings.

In summary, our data suggest that plasma p-tau217 and Tau-PET show similar associations with future cognitive decline and clinical progression in a CU population. We also showed that plasma p-tau217 and Tau-PET provide complimentary information and that a two-step approach (i.e., plasma p-tau217 followed by Tau-PET) substantially reduces the number of required Tau-PET scans and screened participants. Altogether, our data support the feasibility of a clinical trial design in which all participants undergo screening with plasma p-tau217, but only a subset with high/abnormal plasma p-tau217 will undergo Tau-PET.

## CONTRIBUTORS

RO, GS and OH designed the study. RO and GS had full access to raw data and carried out the statistical analyses. RO, GS and OH wrote the manuscript and had the final responsibility to submit for publication. All other authors contributed demographic, clinical, biomarker and neuroimaging data, contributed to the interpretation of the results and critically reviewed the manuscript.

## DATA AVAILABILITY

Due to the multicenter design of the study, access to individual participant data from each cohort will have to be made available through the PIs of the respective cohorts. Generally, anonymised data can be shared by request from qualified academic investigators for the purpose of replicating procedures and results presented in the article, if data transfer is in agreement with the data protection regulation at the institution and is approved by the local Ethics Review Board.

## CODE AVAILABILITY

The codes used for data analyses in our study can be requested from the corresponding authors (RO or OH).

## METHODS

### Participants

We included 1534 participants from the Swedish BioFINDER-1 (n=39, NCT01208675) and BioFINDER-2 (n=481, NCT03174938) studies at Lund University^7,38^, the Mayo Clinic Olmsted Study of Aging^39^ (MCSA, n= n=363), the Australian Imaging Biomarkers and Lifestyle Study of Ageing^40^ (AIBL, n=180), the Knight ADRC at Washington University (n=109), the Translational Biomarkers in Aging and Dementia (TRIAD, n=124) and the PRe-symptomatic EValution of Experimental or Novel Treatments for Alzheimer’s Disease (PREVENT-AD, n=112) at McGill University, the Wisconsin Registry for Alzheimer’s Prevention (WRAP, n=82) at the University of Wisconsin-Madison, and the SCIENCe project^41^, which is part of the Amsterdam Dementia Cohort (ADC, n=44). A brief description of each cohort is provided in **Extended-Data Table-12.** All participants were i) cognitively unimpaired at baseline defined by neuropsychological test scores within the normative range given an individuals’ age, sex and educational background, ii) had Amyloid-PET available to determine Aβ status, iii) underwent a Tau-PET scan and blood sampling within a maximum 1 year interval, and iv) had at least one clinical follow-up visit available. Follow-up data was collected until November 1^st^, 2023. Written informed consent was obtained from all participants and local institutional review boards for human research approved the study.

### Amyloid-PET status

Aβ status was determined using center-specific cut-offs or visual read metrics using [^18^F]flutemetamol PET for BioFINDER-1 and BioFINDER-2, [^11^C]Pittsburgh compound-B PET for MCSA, Knight ADRC and WRAP, [^18^F]florbetapir PET for ADC, and [^18^F]NAV4694 for TRIAD, PREVENT-AD and AIBL, see **Extended-Data Table-13** for details.

### Tau PET

Tau-PET was performed using [^18^F]flortaucipir for BioFINDER-1, MSCA, Knight ADRC, and ADC cohorts, using [^18^F]MK6240 for TRIAD, AIBL and WRAP and using [^18^F]RO948 for BioFINDER-2. Data were centrally processed at Lund University for BioFINDER-1, BioFINDER-2, and ADC, and locally processed for the other cohorts, following previously described procedures (see **Extended-Data Table-14**). In line with our previous study^34^, we generated Tau-PET standardized uptake value ratios (SUVR) for a medial temporal lobe (MTL; unweighted average of bilateral entorhinal cortex and amygdala) and a neocortical temporal (NEO; weighted average of bilateral middle temporal and inferior temporal gyri) region-of-interest.^3,42^

### Plasma p-tau217

Plasma p-tau217 levels were measured using an immunoassay developed by Lilly Research Laboratories (IN, USA) on a Meso Scale Discovery platform at Lund University for BioFINDER-1, BioFINDER-2, ADC, WRAP, PREVENT-AD and Knight ADRC^43^ and at the Mayo Clinic for MSCA^15^. For AIBL and TRIAD a plasma p-tau217+ assay developed by Janssen R&D (CA, USA) on a Single Molecule Array (Simoa) HD-X platform was used.^22^ The correspondence between the two plasma p-tau217 assays used in this study have been shown to be high in a direct head-to-head comparisons study.^44^

### Clinical outcome measures

We used both continuous and binary measures of clinical progression. First, we examined cognitive trajectories using a sensitive composite measure specifically developed to detect cognitive changes in preclinical stages of AD (i.e., the modified preclinical Alzheimer cognitive composite 5 [mPACC5^25,45^]). The mPACC5 consists of tests capturing episodic memory, executive function, semantic memory and global cognition.^25^ Individual neuropsychological tests were z-transformed using the baseline test scores of Aβ-negative participants in each cohort as reference group and then averaged to obtain a composite z-score. The composition of the mPACC5 for each cohort is described in **Extended-Data Table-15**. Second, we examined progression from CU to MCI. MCI was established using the Petersen criteria^46^ and is defined as significant cognitive symptoms as assessed by a physician, in combination with cognitive impairment on one or multiple domains (e.g., memory, executive functioning, attention, language) that is below the normative range given an individuals’ age, sex and educational background, but not sufficiently severe to meet diagnostic criteria for dementia.^47^

### Statistical analyses

All statistical analyses were performed in R version 4.3.1. Statistical significance for all models was set at p<0.05 two-sided, without correction for multiple comparisons. To enable pooling of the data across cohorts, we z-transformed both Tau-PET SUVR’s and plasma p-tau217 concentrations based on the mean and the standard deviation of the Aβ negative participants within each cohort. We conducted two sets of main analyses, in which we examined i) the individual and combined utility of the tau biomarkers for their associatons with longitudinal cognitive decline on the mPACC5 and with clinical progression from CU to MCI, and ii) a sequential two-step approach for participant selection in a preclinical AD trial with a cognitive endpoint.

#### Tau-PET and plasma p-tau217 vs mPACC5 change and progression to MCI

Within these analyses, we performed three different statistical models, including i) only plasma p-tau217 concentration as a (continuous) predictor, ii) only a Tau-PET measure as (continuous) predictor (i.e., Tau-PET_MTL_ or Tau-PET_NEO_ SUVR), or iii) including plasma p-tau217 and one of the Tau-PET measures (i.e., Tau-PET_MTL_ or Tau-PET_NEO_ SUVR) simultaneously as predictors. Additionally, we tested two basic models, model 1 including age, sex, years of education and cohort, and model 2 consisting of the model 1 variables and *APOE* ε4 carriership. We calculated change in mPACC5 using linear mixed models with random time slopes and random intercept using the *lme4* package. Subsequently, these slopes were entered as the dependent variable in linear regression models with plasma p-tau217 and/or Tau-PET as predictors, while adjusting for age, sex, education, cohort and *APOE* ε4 carriership. Differences in model performance was assessed by comparing differences in R^2^ by bootstrapping (1,000 repetitions with resample, *boot* package). Partial R^2^ were calculated to assess the specific contribution of each predictor in the combined plasma p-tau217 and Tau-PET models (*rsq* package).

Next, we examined progression from cognitively unimpaired to MCI using Cox proportional hazard models, adjusting for age, sex, years of education, *APOE*-ε4 carriership and cohort using the continuous variables as predictors. For individuals who progressed to MCI we used the respective times at diagnostic progression to MCI for the analyses. To compare the predictive value of Tau-PET vs plasma p-tau217 for diagnostic conversion, we compared the difference in AICc using bootstrapping procedures (1,000 repetitions with resample). In secondary analyses, all the above-mentioned analyses were repeated in Aβ positive participants.

To assess potential differences between cohorts we performed several sensitivity analyses. First, for the associations between the tau biomarkers and mPACC5 decline we examined the effect sizes and total variance explained in all the previous models for each cohort independently (**Extended-Data Figures-2/3**). To that end, we applied the same linear regression models used in the main analyses to each cohort separately, without including cohort as a covariate. Similarly, we performed Cox proportional hazards models in all cohorts independently to calculate the hazard ratios and C-indices (**Extended-Data Figures-4/5**). Finally, we calculated the root mean square error (RMSE) of the predictions for each cohort when it was excluded from the training set. To do this, we calculated the cognition slopes for all cohorts combined, regressed out the cohort variable using a linear regression model, and then trained a model (with age, sex, and *APOE* as covariates) on all cohorts except one, subsequently predicting cognitive decline in the excluded cohort. We then calculated the RMSE from the difference between the predicted vs the observed data.

#### A two-step approach for participant selection in a preclinical AD trial

To derive optimal sample size reduction for a clinical trial, we generated a data-driven estimate of the complementary value of Tau-PET and plasma p-tau217 when implementing a sequential two-step approach (i.e., plasma p-tau217 first, followed by Tau-PET). First, we calculated the obtained sample size reduction when assuming 80% power to detect a 30% change in cognitive change (mPACC5) in a 4-year clinical trial (with annual repeated testing). Sample size was then defined by using different percentiles (75th, 50th and 25th) of the participants’ baseline plasma p-tau217 levels using the *lmmpower* function in the *longpower* package. The approach was repeated selecting the 75th, 50th and 25th percentiles of the new participants’ Tau-PET measures. Percentage of participants needed compared to the entire study population and the plasma-selected sample are reported. Similar analyses were performed for progression to MCI, using the *ssizeCT.default* function from the *powerSurvEpi* package. In this analysis, we aimed to detect a 30% reduction of events (i.e., progression to MCI) at 80% power.

Next, we compared the characteristics of the sample included and excluded from the hypothetical clinical trial based on the different approaches presented. We focused on four combinations based on the participants selected on their plasma p-tau217 (step 1) and the Tau-PET_MTL_ (step 2) levels, i.e., A) a “liberal” group comprising quartile 2 to quartile 4 (Q2-Q4) of plasma p-tau217 levels and Q2-Q4 of Tau-PET of those selected by plasma, B) a “moderate” group consisting of individuals within Q3-Q4 of plasma p-tau217 levels and Q3-Q4 of Tau-PET of those selected by plasma, C) plasma p-tau217 Q4-only, and D) a “conservative” group consisting of individuals within Q4 of plasma p-tau217 levels and Q4 of Tau-PET of those selected by plasma. We compared the proportion of Aβ-positivity, the mPACC slopes, the proportion of progressors to MCI and the final number of participants included based on the selection criteria between these groups. Finally, we investigated the percentage of cost reductions of such approaches for participant selection in a hypothetical clinical trial using either mPACC5 or progression to MCI as the outcome measure assuming 80% power to detect a 30% change mPACC5 or progression to MCI in a 4-year clinical trial (with annual repeated clinical assessments). We provided percentage cost reductions using different ratios of costs for Tau-PET vs plasma p-tau217; 1:5 (i.e., cost of 1 Tau-PET scan resembling 5 plasma p-tau217 assessments), 1:10. 1:15 and 1:20.

