## Supplemental Material for "Prediction of future cognitive decline among cognitively unimpaired individuals using measures of soluble phosphorylated tau or tau tangle pathology"

### EXTENDED DATA CONTENT


| Table/Figure | Title | Page |
|---|---|---|
| Table 1 | Participant characteristics by cohort (all participants) | 2-10 |
| Figure 1 | Association between plasma p-tau217 and Tau-PET across cohorts | 11 |
| Table 2 | Performance indicators of models predicting mPACC5 decline | 12 |
| Table 3 | Comparison of different models predicting mPACC5 decline | 13 |
| Table 4 | Variance explained by different models predicting mPACC5 decline | 14 |
| Figure 2 | Effect sizes for mPACC5 decline by cohort | 15 |
| Figure 3 | Explained variance for mPACC5 decline by cohort | 16 |
| Table 5 | RMSE of different models predicting mPACC5 decline by cohort | 17 |
| Table 6 | Performance of different models predicting clinical progression to MCI | 18 |
| Table 7 | Comparison of different models predicting clinical progression to MCI | 19 |
| Figure 4 | Effect sizes for clinical progression to MCI by cohort | 20 |
| Figure 5 | C-index for clinical progression to MCI by cohort | 21 |
| Table 8 | C-index of different models predicting clinical progression MCI | 22 |
| Figure 6 | Two-step approach for trials using mPACC5 decline with Tau-PET$_{NEO}$ | 23 |
| Figure 7 | Two-step approach for trials using progression to MCI with Tau-PET$_{NEO}$ | 24 |
| Table 9 | Sample size reductions in a clinical trial following a two-step approach | 25 |
| Figure 8 | Two-step approach for trials using mPACC5 decline in Aβ+ CU | 26 |
| Table 10 | Plasma p-tau217 / Tau-PET$_{MTL}$ groups: Aβ status and clinical outcomes | 27 |
| Table 11 | Plasma p-tau217 / Tau-PET$_{MTL}$ groups: Demographic information | 28 |
| Figure 9 | Characterization of different plasma p-tau217/Tau-PET$_{NEO}$ groups | 29 |
| Figure 10 | Saved costs in a hypothetical trial with mPACC5 as an endpoint | 30 |
| Figure 11 | Saved costs in a hypothetical trial with MCI progression as an endpoint | 31 |
| Table 12 | Cohort descriptions | 32-34 |
| Table 13 | Methods to determine Amyloid PET status by cohort | 35-36 |
| Table 14 | Methods to determine Tau PET status | 37 |
| Table 15 | Composition of the mPACC5 for each cohort | 38 |
|  | References | 39 |

**Extended Data Table 1.** Participant characteristics by cohort (all participants)

| ADC | | |
|---|---|---|
| | **All participants** | **Aβ+ participants only** |
| N | 44 | 17 |
| Age, years | 65.0±7.5 | 66.4±6.3 |
| Sex, % female | 45.5% | 47.1% |
| Education, years | 12.1±2.7 | 12.2±2.8 |
| MMSE score | 28.8±1.3 | 28.4±1.3 |
| *APOE e*4 status, % carriers | 38.6% | 64.7% |
| Aβ-status, % positive | 38.6% | 100% |
| Follow-up duration, years | 4.6±1.8 | 3.8±1.6 |
| Follow-up visits, median (range) | 5 (2-8) | 5 (3-7) |
| Plasma p-tau217, z-score | 0.62±1.4 | 1.59±1.28 |
| Tau-$PET_{MTL}$, z-score | 0.71±1.75 | 1.84±2.10 |
| Tau-$PET_{NEO}$, z-score | 0.81±2.50 | 2.10±3.51 |
| mPACC5, baseline score | -0.19±0.74 | -0.50±0.61 |
| mPACC5, annual change | -0.065±0.084 | -0.161±0.148 |
| % Progression to MCI | 13.6% | 35.3% |

| AIBL | | |
|---|---|---|
| | **All participants** | **Aβ+ participants only** |
| N | 180 | 34 |
| Age, years | 74.7±5.3 | 77.5±6.4 |
| Sex, % female | 52.8% | 58.8% |
| Education, years | 12.7±2.7 | 11.5±2.9 |
| MMSE score | 28.5 ±1.4 | 27.9±1.6 |
| *APOE e*4 status, % carriers | 29.4% | 58.8% |
| Aβ-status, % positive | 18.9% | 100% |
| Follow-up duration, years | 3.2±0.8 | 2.9±0.9 |
| Follow-up visits, median (range) | 3 (2-4) | 3 (2-4) |
| Plasma p-tau217, z-score | 0.21±0.99 | 0.95±0.88 |
| Tau-$PET_{MTL}$, z-score | 0.28±1.17 | 1.57±1.30 |
| Tau-$PET_{NEO}$, z-score | 0.27±1.43 | 1.27±2.36 |
| mPACC5, baseline score | -0.02±0.71 | -0.27±0.80 |
| mPACC5, annual change | -0.045±0.068 | -0.130±0.142 |
| % Progression to MCI | 3.9% | 8.8% |

| BioFINDER-1 | | |
|---|---|---|
| | **All participants** | **Aβ+ participants only** |
| N | 39 | 12 |
| Age, years | 73.5±7.0 | 74.2±5.9 |
| Sex, % female | 53.8% | 58.3% |
| Education, years | 11.8±3.7 | 10.7±3.0 |
| MMSE score | 28.6±1.3 | 28.3±1.7 |
| *APOE e*4 status, % carriers | 53.8% | 75.0% |
| Aβ-status, % positive | 30.8% | 100% |
| Follow-up duration, years | 3.4±0.75 | 3.3±0.9 |
| Follow-up visits, median (range) | 2 (2-5) | 2 (2-5) |
| Plasma p-tau217, z-score | 0.16±1.23 | 0.53±1.70 |
| Tau-PET$_{MTL}$, z-score | 0.40±1.69 | 1.40±2.48 |
| Tau-PET$_{NEO}$, z-score | 0.46±1.90 | 1.45±2.99 |
| mPACC5, baseline score | 0.06±0.74 | -0.22±0.87 |
| mPACC5, annual change | -0.037±0.053 | -0.082±0.095 |
| % Progression to MCI | 12.8% | 41.7% |

| BioFINDER-2 | | |
|---|---|---|
| | **All participants** | **Aβ+ participants only** |
| N | 481 | 137 |
| Age, years | 65.0±11.4 | 70.1±9.1 |
| Sex, % female | 52.4% | 49.6% |
| Education, years | 12.8±3.5 | 12.8±3.8 |
| MMSE score | 28.9±1.3 | 28.7±1.4 |
| *APOE e*4 status, % carriers | 48.2% | 71.5% |
| Aβ-status, % positive | 28.5% | 100% |
| Follow-up duration, years | 3.0±1.1 | 3.0±1.2 |
| Follow-up visits, median (range) | 3 (2-6) | 3 (2-6) |
| Plasma p-tau217, z-score | 0.48±1.36 | 1.78±1.23 |
| Tau-$PET_{MTL}$, z-score | 0.26±1.58 | 1.53±2.05 |
| Tau-$PET_{NEO}$, z-score | 0.13±1.66 | 0.99±2.52 |
| mPACC5, baseline score | 0.17±0.78 | -0.11±0.81 |
| mPACC5, annual change | -0.034±0.088 | -0.113±0.174 |
| % Progression to MCI | 11.0% | 26.3% |

| Knight ADRC | | |
|---|---|---|
| | **All participants** | **Aβ+ participants only** |
| N | 109 | 34 |
| Age, years | 70.2±6.4 | 70.6±6.3 |
| Sex, % female | 53.2% | 61.8% |
| Education, years | 16.3±2.3 | 16.6±2.3 |
| MMSE score | 29.3±1.1 | 29.4±1.1 |
| *APOE e*4 status, % carriers | 29.4% | 35.3% |
| Aβ-status, % positive | 31.2% | 100% |
| Follow-up duration, years | 3.9±1.7 | 3.6±1.5 |
| Follow-up visits, median (range) | 4 (2-8) | 4 (2-8) |
| Plasma p-tau217, z-score | 0.71±1.79 | 2.10±2.23 |
| Tau-$PET_{MTL}$, z-score | 0.27±1.21 | 0.85±1.39 |
| Tau-$PET_{NEO}$, z-score | 0.31±1.53 | 0.94±2.17 |
| mPACC5, baseline score | -0.08±0.68 | -0.13±0.76 |
| mPACC5, annual change | -0.050±0.083 | -0.138±0.144 |
| % Progression to MCI | 11.9% | 20.6% |

| MCSA | | |
|---|---|---|
| | **All participants** | **Aβ+ participants only** |
| N | 363 | 108 |
| Age, years | 68.3±12.0) | 76.4±7.9 |
| Sex, % female | 45.7% | 53.7% |
| Education, years | 15.1±2.3 | 14.7±2.5 |
| MMSE score | 28.8±1.0 | 28.5±1.2 |
| *APOE e*4 status, % carriers | 29.2% | 47.2% |
| Aβ-status, % positive | 108 (29.8%) | 100% |
| Follow-up duration, years | 5.6±2.1 | 4.9±2.2 |
| Follow-up visits, median (range) | 5 (2-7) | 5 (2-7) |
| Plasma p-tau217, z-score | 0.42±1.29 | 1.34±1.40 |
| Tau-$PET_{MTL}$, z-score | 0.17±1.18 | 0.76±1.41 |
| Tau-$PET_{NEO}$, z-score | 0.06±1.09 | 0.47±1.20 |
| mPACC5, baseline score | -0.01±0.75 | -0.42±0.67 |
| mPACC5, annual change | -0.038±0.053 | -0.102±0.084 |
| % Progression to MCI | 11.0% | 25.0% |

| PREVENT-AD | | |
|---|---|---|
| | **All participants** | **Aβ+ participants only** |
| N | 112 | 24 |
| Age, years | 67.4±4.8 | 68.5±5.1 |
| Sex, % female | 74.1% | 66.7% |
| Education, years | 15.3±3.31 | 14.3±2.9 |
| MMSE score | 28.8±1.2 | 28.7±1.2 |
| *APOE e*4 status, % carriers | 39.3% | 62.5% |
| Aβ-status, % positive | 21.4% | 100% |
| Follow-up duration, years | 4.2±1.2 | 4.4±1.3 |
| Follow-up visits, median (range) | 4 (2-5) | 3 (2-5) |
| Plasma p-tau217, z-score | 0.34±1.44 | 1.75±1.87 |
| Tau-$PET_{MTL}$, z-score | 0.19±1.14 | 0.93±1.36 |
| Tau-$PET_{NEO}$, z-score | 0.14±1.20 | 0.66±1.71 |
| mPACC5, baseline score | 0.05±0.60 | -0.31±0.60 |
| mPACC5, annual change | -0.021±0.061 | -0.058±0.135 |
| % Progression to MCI | 22.3% | 54.2% |

| TRIAD | | |
|---|---|---|
| | **All participants** | **Aβ+ participants only** |
| N | 124 | 27 |
| Age, years | 71.4±5.8 | 74.2±4.8 |
| Sex, % female | 66.9% | 74.1% |
| Education, years | 15.7±3.6 | 14.1±3.2 |
| MMSE score | 29.2±0.9 | 29.0±1.1 |
| *APOE e*4 status, % carriers | 22.6% | 25.9% |
| Aβ-status, % positive | 21.8% | 100% |
| Follow-up duration, years | 2.4±0.7 | 2.2±0.5 |
| Follow-up visits, median (range) | 3 (2-4) | 3 (2-4) |
| Plasma p-tau217, z-score | 0.31±1.20 | 1.61±0.98 |
| Tau-$PET_{MTL}$, z-score | 0.36±1.38 | 1.55±1.88 |
| Tau-$PET_{NEO}$, z-score | 0.15±1.12 | 0.60±1.28 |
| mPACC5, baseline score | -0.02±0.75 | -0.083±0.81 |
| mPACC5, annual change | -0.053±0.070 | -0.107±0.160 |
| % Progression to MCI | 13.7% | 33.3% |

| WRAP | | |
|---|---|---|
| | **All participants** | **Aβ+ participants only** |
| N | 82 | 20 |
| Age, years | 68.1±5.9 | 70.5±4.5 |
| Sex, % female | 58.5% | 50.0% |
| Education, years | 16.5±2.1 | 17.1±2.1 |
| MMSE score | 29.4±0.9 | 28.9±1.3 |
| *APOE e*4 status, % carriers | 41.5% | 55.0% |
| Aβ-status, % positive | 24.4% | 100% |
| Follow-up duration, years | 3.0±1.1 | 2.68±0.79 |
| Follow-up visits, median (range) | 2 (2-3) | 2 (2-3) |
| Plasma p-tau217, z-score | 0.70±1.66 | 2.82±1.43 |
| Tau-$PET_{MTL}$, z-score | 0.43±1.79 | 1.90±2.66 |
| Tau-$PET_{NEO}$, z-score | 0.25±1.53 | 0.93±2.52 |
| mPACC5, baseline score | 0.01±0.74 | -0.22±0.88 |
| mPACC5, annual change | -0.053±0.083 | -0.121±0.140 |
| % Progression to MCI | 7.3% | 25.0% |

**Extended Data Figure 1.** The association between plasma p-tau217 and Tau-PET$_{MTL}$/Tau-PET$_{NEO}$ across cohorts

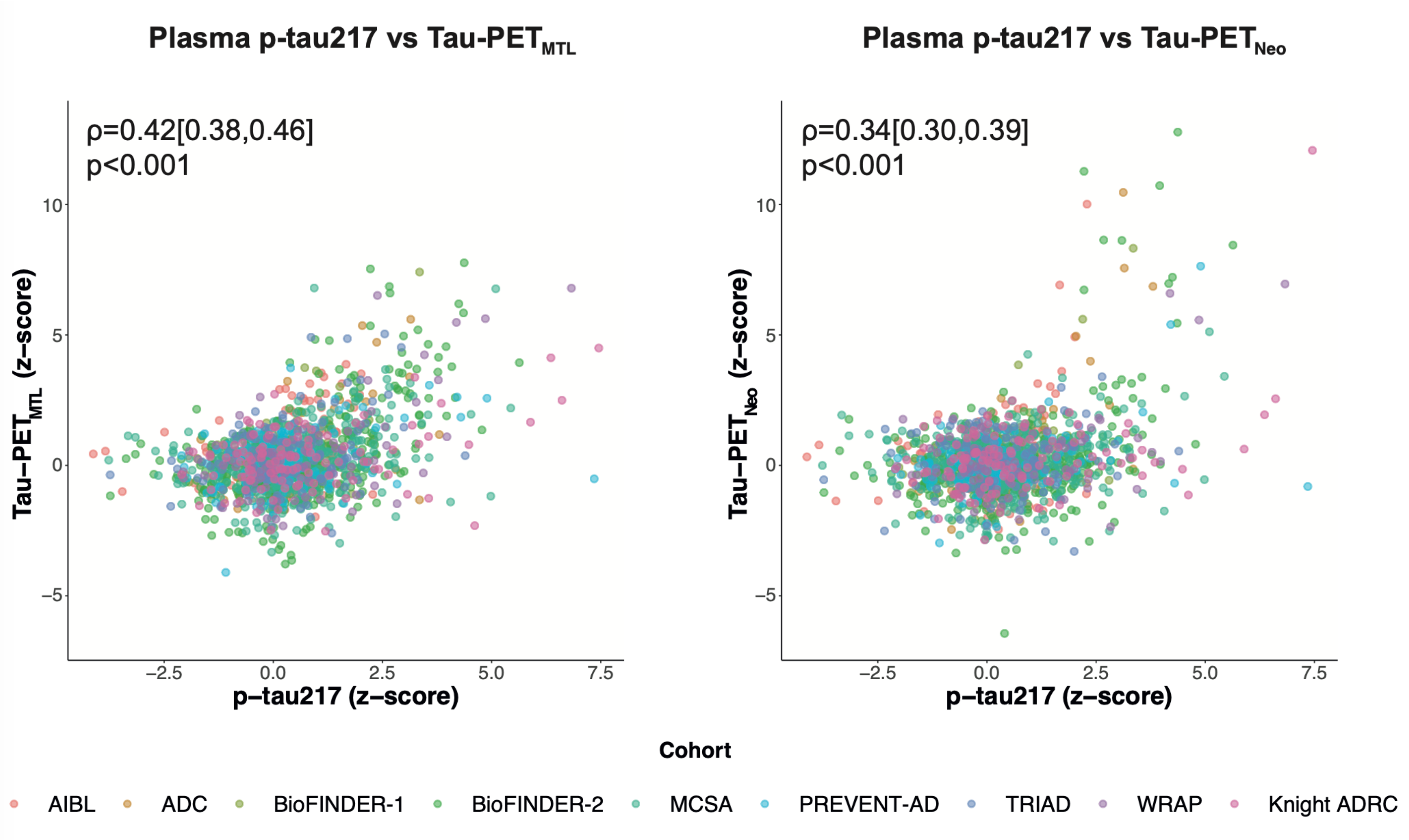


Spearman correlations are presented, color coded by cohort.

**Extended Data Table 2.** Performance indicators of models predicting decline on the mPACC5

| **Model** | **plasma p-tau217 $\beta_{std}$ [95%CI]** | **p plasma p-tau217** | **Tau-PET $\beta_{std}$ [95%CI]** | **p Tau-PET** | **$R^2$** | **AICc** |
|---|---|---|---|---|---|---|
| **All participants** | | | | | | |
| Basic without *APOE* | - | - | - | - | 0.23[0.19, 0.26] | -3603.3 |
| Basic with *APOE* | - | - | - | - | 0.24[0.20, 0.27] | -3617.1 |
| Plasma p-tau217 | -0.02 [-0.02, -0.01] | <0.001 | - | - | 0.32[0.27, 0.35] | -3766.1 |
| Tau-PET MTL | - | - | -0.02 [-0.02, -0.01] | <0.001 | 0.32[0.27, 0.36] | -3773.5 |
| Tau-PET Neo-T | - | - | -0.01 [-0.02, -0.01] | <0.001 | 0.31[0.25, 0.35] | -3750.9 |
| Plasma p-tau217 & Tau-PET MTL | -0.01 [-0.02, -0.01] | <0.001 | -0.01 [-0.02, -0.01] | <0.001 | 0.36[0.30, 0.4] | -3848.9 |
| Plasma p-tau217 & Tau-PET Neo-T | -0.01 [-0.02, -0.01] | <0.001 | -0.01 [-0.01, -0.01] | <0.001 | 0.35[0.29, 0.4] | -3841.1 |
| **Aβ+ participants** | | | | | | |
| Basic without *APOE* | - | - | - | - | 0.16[0.07, 0.21] | -427.7 |
| Basic with *APOE* | - | - | - | - | 0.16[0.07, 0.21] | -427.6 |
| Plasma p-tau217 | -0.04 [-0.05, -0.03] | <0.001 | - | - | 0.30[0.19, 0.36] | -497.2 |
| Tau-PET MTL | - | - | -0.04 [-0.04, -0.03] | <0.001 | 0.33[0.22, 0.40] | -515.0 |
| Tau-PET Neo-T | - | - | -0.03 [-0.04, -0.02] | <0.001 | 0.35[0.22, 0.43] | -523.2 |
| Plasma p-tau217 & Tau-PET MTL | -0.03 [-0.04, -0.02] | <0.001 | -0.03 [-0.04, -0.02] | <0.001 | 0.38[0.27, 0.45] | -545.5 |
| Plasma p-tau217 & Tau-PET Neo-T | -0.03 [-0.04, -0.02] | <0.001 | -0.02 [-0.03, -0.02] | <0.001 | 0.39[0.27, 0.47] | -550.4 |

**Extended Data Table 3.** Comparison of different models predicting cognitive decline on the mPACC5

| **P-values** | Basic without APOE | Basic with APOE | Plasma p-tau217 | Tau-PET$_{MTL}$ | Tau-PET$_{NEO}$ | Plasma p-tau217 & Tau-PET$_{MTL}$ | Plasma p-tau217 & Tau-PET$_{NEO}$ |
|---|---|---|---|---|---|---|---|
| **All Participants** | | | | | | | |
| Basic without *APOE* | 1 | 0.054 | <0.001 | <0.001 | <0.001 | <0.001 | <0.001 |
| Basic with *APOE* | | 1 | <0.001 | <0.001 | 0.001 | <0.001 | <0.001 |
| Plasma p-tau217 | | | 1 | 0.812 | 0.699 | <0.001 | 0.004 |
| Tau-PET MTL | | | | 1 | 0.404 | <0.001 | 0.019 |
| Tau-PET Neo-T | | | | | 1 | 0.002 | <0.001 |
| Plasma p-tau217 & Tau-PET MTL | | | | | | 1 | 0.713 |
| Plasma p-tau217 & Tau-PET Neo-T | | | | | | | 1 |
| **Aβ+ participants** | | | | | | | |
| Basic without *APOE* | 1 | 0.750 | <0.001 | <0.001 | <0.001 | <0.001 | <0.001 |
| Basic with *APOE* | | 1 | <0.001 | <0.001 | <0.001 | <0.001 | <0.001 |
| Plasma p-tau217 | | | 1 | 0.344 | 0.287 | 0.003 | 0.001 |
| Tau-PET MTL | | | | 1 | 0.693 | 0.002 | 0.051 |
| Tau-PET Neo-T | | | | | 1 | 0.313 | 0.008 |
| Plasma p-tau217 & Tau-PET MTL | | | | | | 1 | 0.760 |
| Plasma p-tau217 & Tau-PET Neo-T | | | | | | | 1 |

**Extended Data Table 4.** Variance explained by different models predicting cognitive decline on the mPACC5

| **Model** | **Total $R^2$** | **Partial $R^2$ covariates** | **Partial $R^2$ plasma p-tau217** | **Partial $R^2$ Tau-PET** | **Partial $R^2$ shared** |
|---|---|---|---|---|---|
| **All participants** | | | | | |
| Basic without *APOE* | 0.23 | 0.25 | - | - | 0.00 |
| Basic with *APOE* | 0.24 | 0.26 | - | - | 0.00 |
| Plasma p-tau217 | 0.32 | 0.19 | 0.1 | - | 0.03 |
| Tau-PET MTL | 0.32 | 0.19 | - | 0.1 | 0.02 |
| Tau-PET Neo-T | 0.31 | 0.23 | - | 0.09 | 0.00 |
| Plasma p-tau217 & Tau-PET MTL | 0.36 | 0.16 | 0.05 | 0.06 | 0.08 |
| Plasma p-tau217 & Tau-PET Neo-T | 0.35 | 0.19 | 0.06 | 0.05 | 0.05 |
| **Aβ+ participants** | | | | | |
| Basic without *APOE* | 0.16 | 0.19 | - | - | 0.00 |
| Basic with *APOE* | 0.16 | 0.19 | - | - | 0.00 |
| Plasma p-tau217 | 0.30 | 0.10 | 0.16 | - | 0.03 |
| Tau-PET MTL | 0.33 | 0.14 | - | 0.20 | 0.00 |
| Tau-PET Neo-T | 0.35 | 0.18 | - | 0.22 | 0.00 |
| Plasma p-tau217 & Tau-PET MTL | 0.38 | 0.1 | 0.08 | 0.12 | 0.09 |
| Plasma p-tau217 & Tau-PET Neo-T | 0.39 | 0.11 | 0.07 | 0.13 | 0.08 |

**Extended Data Figure 2.** Effect sizes for mPACC5 decline by cohort

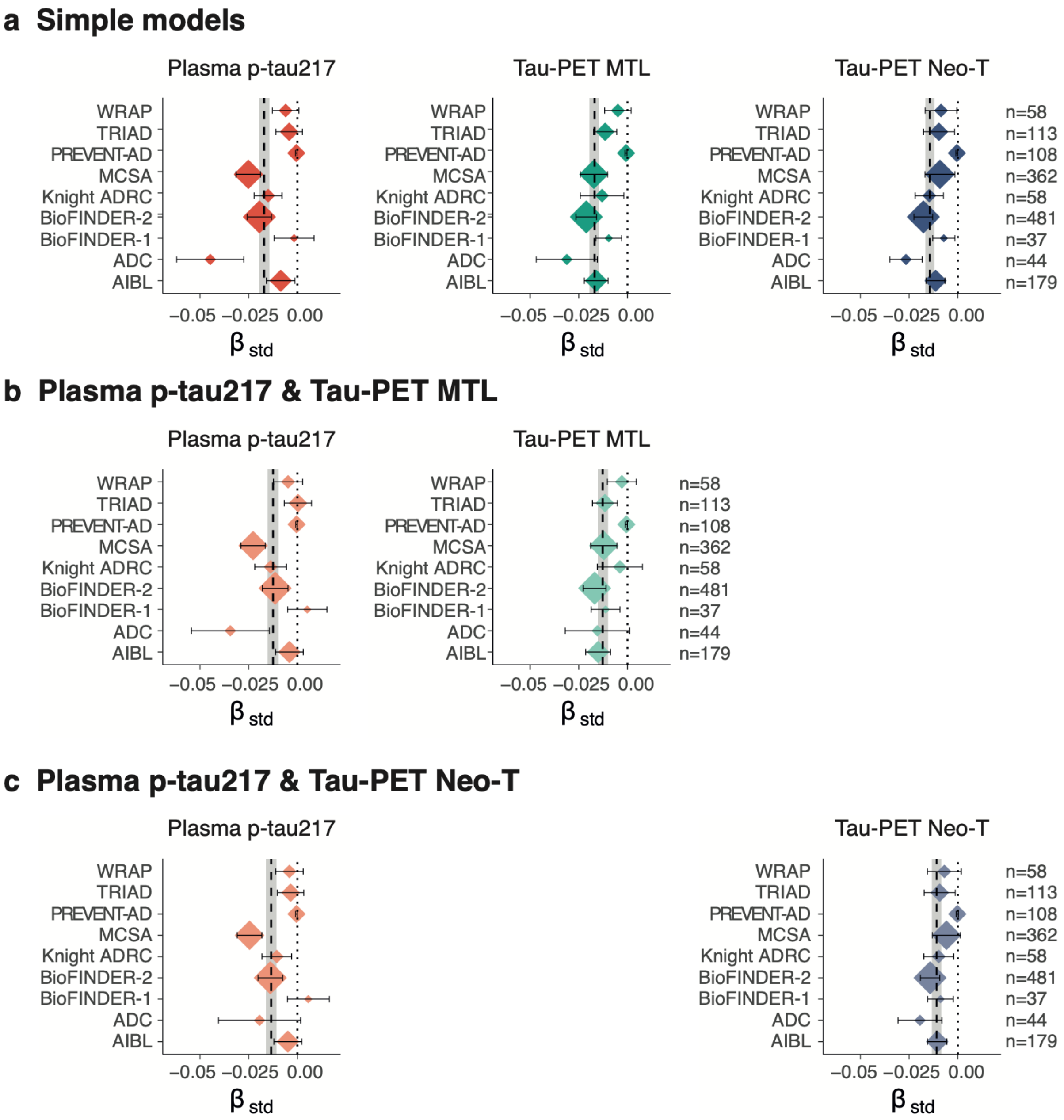


Effect sizes (expressed as standardized beta's) for predicting longitudinal changes on the mPACC5 in each of the cohorts. The vertical dotted line represents standardized beta = 0, while the vertical dashed line represent the average standardized beta across all cohorts with the 95% CI indicated in gray. Errorbars represent the 95%CI for each cohort. The size of the diamonds are proportional to the sample size of each cohort. Panel **a** shows the individual tau biomarker models, while **b,c** show combined models of plasma p-tau217 and Tau-PET.

**Extended Data Figure 3.** Explained variance for mPACC5 decline by cohort

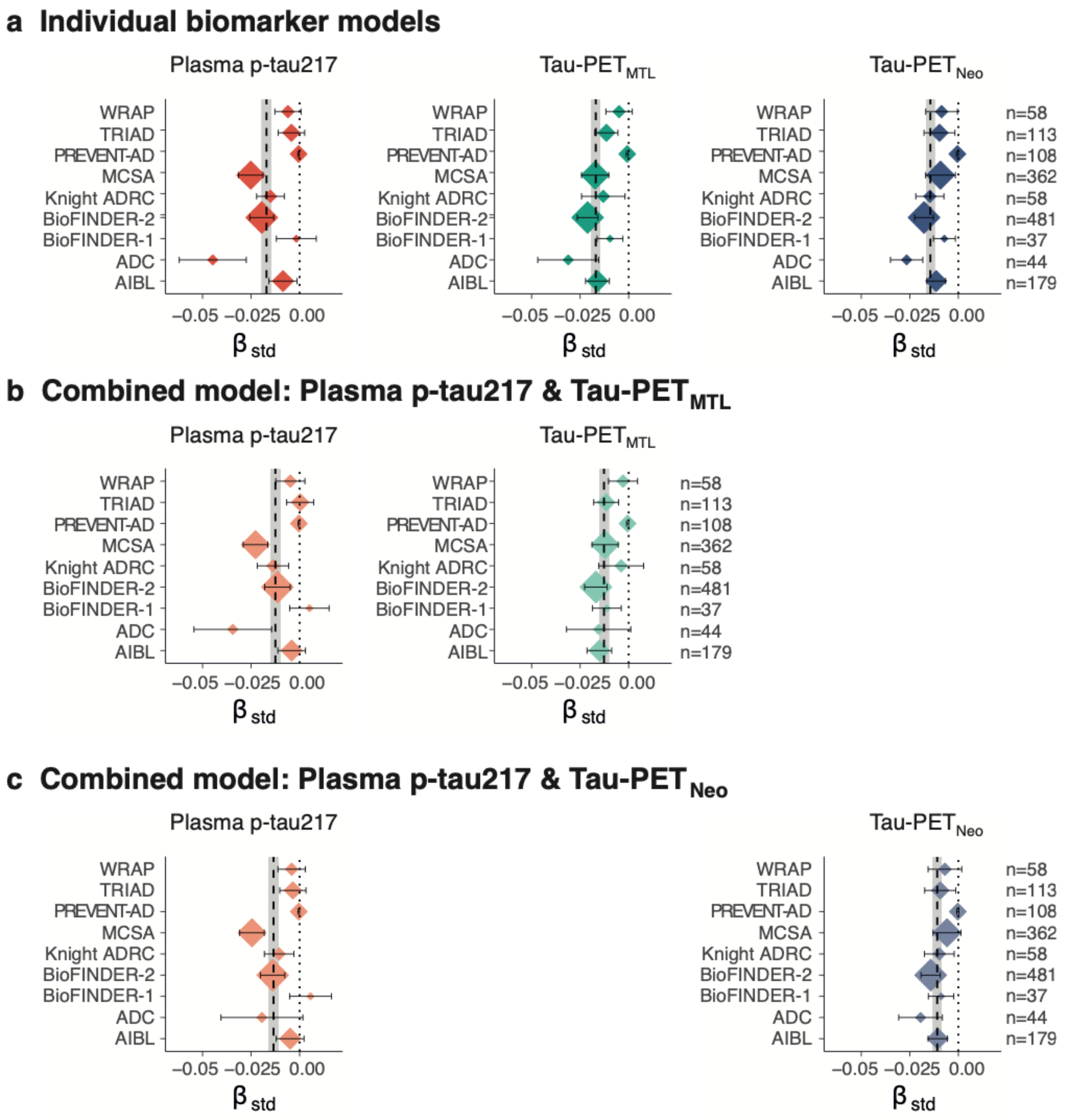


Explained variance (expressed as $R^2$) for predicting longitudinal changes on the mPACC5 in each of the cohorts. The vertical dotted line represents $R^2 = 0$, while the vertical dashed line represent the average $R^2$ across all cohorts with the 95%CI indicated in gray. Errorbars represent the 95% CI for each cohort. The size of the diamonds are proportional to the sample size of each cohort. Panel **a** shows the individual tau biomarker models, while **b** shows combined models of plasma p-tau217 and Tau-PET.

**Extended Data Table 5.** Performance indicator (RMSE) of different models predicting decline on the mPACC5 by cohort

| Cohort | N | Basic without APOE | Basic with APOE | Plasma p-tau217 | Tau-PET$_{MTL}$ | Tau-PET $_{NEO}$ | Plasma p-tau217 & Tau-PET$_{MTL}$ | Plasma p-tau217 & Tau-PET$_{NEO}$ |
|---|---|---|---|---|---|---|---|---|
| ADC | 44 | 0.078 [0.078,0.078] | 0.076 [0.075,0.077] | 0.065 [0.062,0.067] | 0.065 [0.063,0.066] | 0.059 [0.055,0.061] | 0.060 [0.058,0.061] | 0.056 [0.053,0.058] |
| AIBL | 179 | 0.066 [0.064,0.068] | 0.066 [0.064,0.067] | 0.063 [0.062,0.064] | 0.061 [0.059,0.062] | 0.063 [0.061,0.064] | 0.060 [0.058,0.061] | 0.061 [0.059,0.062] |
| BioFINDER-1 | 37 | 0.048 [0.045,0.051] | 0.048 [0.045,0.051] | 0.047 [0.044,0.049] | 0.043 [0.039,0.046] | 0.046 [0.042,0.050] | 0.044 [0.041,0.047] | 0.047 [0.043,0.050] |
| BioFINDER-2 | 481 | 0.078 [0.077,0.079] | 0.077 [0.076,0.078] | 0.073 [0.072,0.074] | 0.072 [0.071,0.073] | 0.073 [0.072,0.074] | 0.071 [0.069,0.071] | 0.071 [0.070,0.072] |
| Knight ADRC | 58 | 0.077 [0.076,0.077] | 0.076 [0.075,0.077] | 0.064 [0.063,0.065] | 0.072 [0.071,0.072] | 0.067 [0.066,0.068] | 0.065 [0.063,0.065] | 0.061 [0.060,0.062] |
| MCSA | 362 | 0.072 [0.071,0.073] | 0.071 [0.070,0.072] | 0.066 [0.065,0.067] | 0.071 [0.070,0.071] | 0.069 [0.068,0.070] | 0.067 [0.066,0.068] | 0.066 [0.065,0.066] |
| PREVENT-AD | 108 | 0.049 [0.047,0.050] | 0.049 [0.047,0.050] | 0.047 [0.046,0.049] | 0.049 [0.047,0.050] | 0.047 [0.045,0.048] | 0.048 [0.046,0.049] | 0.046 [0.044,0.047] |
| TRIAD | 113 | 0.057 [0.056,0.057] | 0.058 [0.057,0.058] | 0.061 [0.059,0.062] | 0.056 [0.055,0.057] | 0.056 [0.055,0.056] | 0.059 [0.057,0.060] | 0.059 [0.057,0.060] |
| WRAP | 58 | 0.068 [0.067,0.069] | 0.068 [0.067,0.069] | 0.065 [0.063,0.066] | 0.062 [0.060,0.063] | 0.063 [0.061,0.064] | 0.061 [0.059,0.062] | 0.062 [0.060,0.063] |

RMSE = Root-mean-square deviation

**Extended Data Table 6.** Performance of different models predicting clinical progression to mild cognitive impairment (MCI)

| Model | N non-progressor | N progressor | HR plasma p-tau217 | p plasma p-tau217 | HR Tau-PET | p Tau-PET | C-index | AICc |
|---|---|---|---|---|---|---|---|---|
| **All participants** | | | | | | | | |
| Basic without *APOE* | 1320 | 172 | | - | | - | 0.75 | 2205 |
| Basic with *APOE* | 1320 | 172 | | - | | - | 0.76 | 2185 |
| Plasma p-tau217 | 1320 | 172 | 1.57 [1.44, 1.71] | <0.001 | | - | 0.82 | 2099 |
| Tau-PET$_{MTL}$ | 1320 | 172 | | - | 1.63 [1.50, 1.77] | <0.001 | 0.82 | 2077 |
| Tau-PET$_{NEO}$ | 1320 | 172 | | - | 1.42 [1.33, 1.51] | <0.001 | 0.81 | 2111 |
| Plasma p-tau217 & Tau-PET$_{MTL}$ | 1320 | 172 | 1.37 [1.24, 1.52] | <0.001 | 1.43 [1.30, 1.57] | <0.001 | 0.84 | 2047 |
| Plasma p-tau217 & Tau-PET$_{NEO}$ | 1320 | 172 | 1.42 [1.29, 1.57] | <0.001 | 1.25 [1.16, 1.34] | <0.001 | 0.83 | 2069 |
| **Aβ+ participants** | | | | | | | | |
| Basic without *APOE* | 292 | 111 | | - | | - | 0.66 | 1177 |
| Basic with *APOE* | 292 | 111 | | - | | - | 0.67 | 1175 |
| Plasma p-tau217 | 292 | 111 | 1.56 [1.37, 1.77] | <0.001 | | - | 0.75 | 1133 |
| Tau-PET$_{MTL}$ | 292 | 111 | | - | 1.54 [1.39, 1.70] | <0.001 | 0.77 | 1109 |
| Tau-PET$_{NEO}$ | 292 | 111 | | - | 1.34 [1.25, 1.43] | <0.001 | 0.74 | 1126 |
| Plasma p-tau217 & Tau-PET$_{MTL}$ | 292 | 111 | 1.39 [1.21, 1.60] | <0.001 | 1.42 [1.28, 1.58] | <0.001 | 0.78 | 1092 |
| Plasma p-tau217 & Tau-PET$_{NEO}$ | 292 | 111 | 1.40 [1.21, 1.61] | <0.001 | 1.24 [1.15, 1.33] | <0.001 | 0.77 | 1108 |

**Extended Data Table 7.** Comparison (p-values) of different models predicting clinical progression to mild cognitive impairment (MCI)

| **P-values** | Basic without APOE | Basic with APOE | Plasma p-tau217 | Tau-$PET_{MTL}$ | Tau-$PET_{NEO}$ | Plasma p-tau217 & Tau-$PET_{MTL}$ | Plasma p-tau217 & Tau-$PET_{NEO}$ |
|---|---|---|---|---|---|---|---|
| **All Participants** | | | | | | | |
| Basic without APOE | 1 | 0.025 | <0.001 | <0.001 | <0.001 | <0.001 | <0.001 |
| Basic with APOE | | 1 | <0.001 | <0.001 | 0,001 | <0.001 | <0.001 |
| Plasma p-tau217 | | | 1 | 0.34 | 0.571 | 0.005 | 0.018 |
| Tau-$PET_{MTL}$ | | | | 1 | 0.046 | 0.007 | 0.682 |
| Tau-$PET_{NEO}$ | | | | | 1 | 0.001 | 0.001 |
| Plasma p-tau217 & Tau-$PET_{MTL}$ | | | | | | 1 | 0.072 |
| Plasma p-tau217 & Tau-$PET_{NEO}$ | | | | | | | 1 |
| **Aβ+ participants** | | | | | | | |
| Basic without APOE | 1 | 0.621 | 0,01 | <0.001 | 0,003 | <0.001 | <0.001 |
| Basic with APOE | | 1 | 0.012 | <0.001 | 0,002 | <0.001 | <0.001 |
| Plasma p-tau217 | | | 1 | 0.186 | 0.721 | 0.002 | 0.03 |
| Tau-$PET_{MTL}$ | | | | 1 | 0.177 | 0.043 | 0.923 |
| Tau-$PET_{NEO}$ | | | | | 1 | 0.023 | 0.049 |
| Plasma p-tau217 & Tau-$PET_{MTL}$ | | | | | | 1 | 0.099 |
| Plasma p-tau217 & Tau-$PET_{NEO}$ | | | | | | | 1 |

**Extended Data Figure 4.** Effect sizes for clinical progression to MCI by cohort

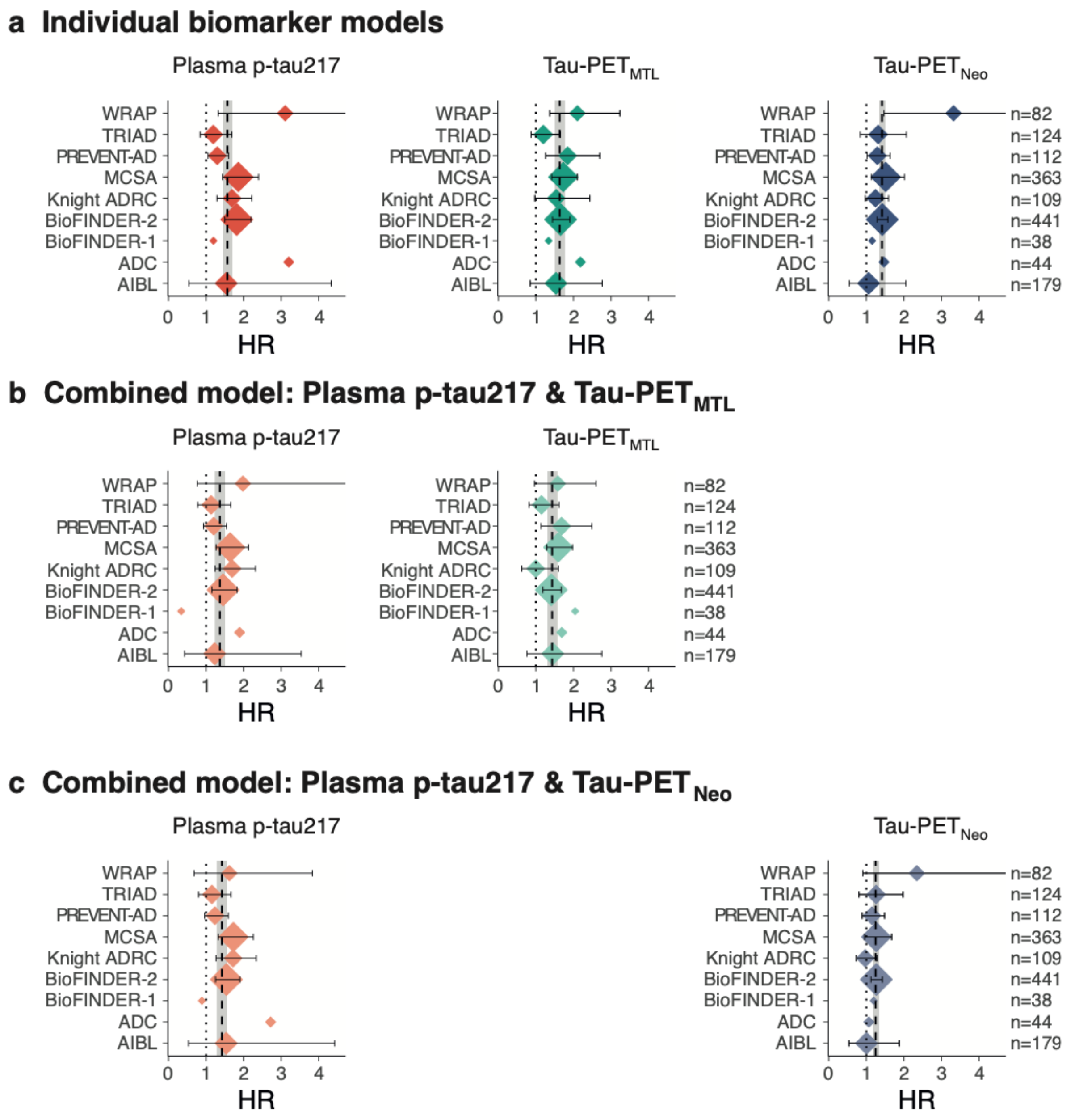


Effect sizes (expressed as hazard ratios [HR]) for predicting future clinical progression to mild cognitive impairment in each of the cohorts. The vertical dotted line represents HR = 1, while the vertical dashed line represent the average HR across all cohorts with the 95% CI indicated in gray. Errorbars represent the 95%CI for each cohort. The size of the diamonds are proportional to the sample size of each cohort. Panel **a** shows the individual tau biomarker models, while **b,c** show combined models of plasma p-tau217 and Tau-PET.

**Extended Data Figure 5.** C-index for clinical progression to MCI by cohort

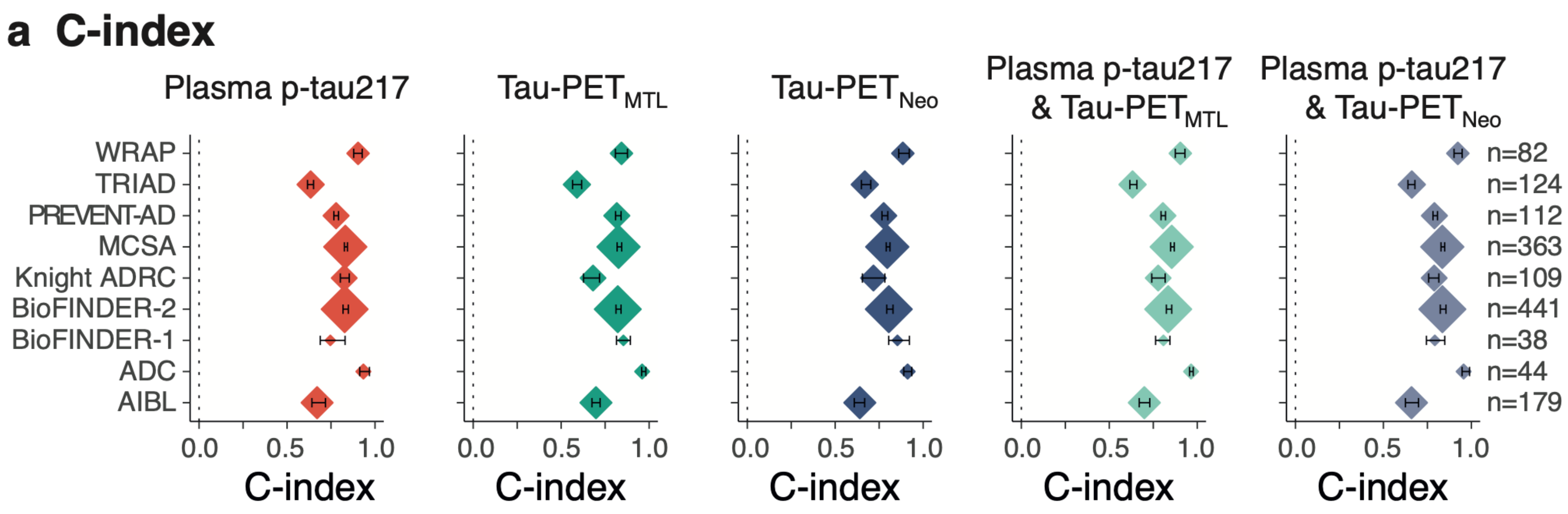


Model fit (expressed as the C-index) for predicting future clinical progression to mild cognitive impairment in each of the cohorts. The vertical dotted line represents C-index = 0. Errorbars represent the 95%CI for each cohort. The size of the diamonds are proportional to the sample size of each cohort.

**Extended Data Table 8.** C-index of different models predicting clinical progression to mild cognitive impairment (MCI)

| Cohort | N | Basic without APOE | Basic with APOE | Plasma p-tau217 | Tau-PET$_{MTL}$ | Tau-PET$_{NEO}$ | Plasma p-tau217 & Tau-PET$_{MTL}$ | Plasma p-tau217 & Tau-PET$_{NEO}$ |
|---|---|---|---|---|---|---|---|---|
| ADC | 44 | 0.689 [0.636,0.747] | 0.804 [0.772,0.874] | 0.934 [0.913,0.968] | 0.912 [0.890,0.935] | 0.960 [0.958,0.982] | 0.956 [0.947,0.990] | 0.965 [0.956,0.977] |
| AIBL | 179 | 0.622 [0.573,0.670] | 0.635 [0.595,0.674] | 0.671 [0.641,0.718] | 0.640 [0.608,0.668] | 0.698 [0.675,0.721] | 0.660 [0.625,0.700] | 0.699 [0.670,0.730] |
| BioFINDER-1 | 38 | 0.612 [0.527,0.712] | 0.762 [0.660,0.852] | 0.746 [0.689,0.830] | 0.854 [0.804,0.922] | 0.854 [0.815,0.893] | 0.792 [0.745,0.849] | 0.808 [0.763,0.845] |
| BioFINDER-2 | 441 | 0.711 [0.700,0.722] | 0.729 [0.720,0.745] | 0.828 [0.818,0.848] | 0.805 [0.793,0.829] | 0.822 [0.812,0.840] | 0.835 [0.823,0.855] | 0.835 [0.824,0.855] |
| Knight ADRC | 109 | 0.808 [0.771,0.845] | 0.739 [0.696,0.791] | 0.825 [0.803,0.854] | 0.718 [0.655,0.782] | 0.681 [0.627,0.718] | 0.789 [0.757,0.815] | 0.778 [0.741,0.818] |
| MCSA | 363 | 0.749 [0.740,0.759] | 0.772 [0.760,0.789] | 0.831 [0.826,0.843] | 0.796 [0.790,0.811] | 0.826 [0.818,0.844] | 0.835 [0.828,0.848] | 0.855 [0.849,0.869] |
| PREVENT-AD | 112 | 0.705 [0.678,0.744] | 0.727 [0.719,0.752] | 0.779 [0.766,0.793] | 0.776 [0.766,0.799] | 0.816 [0.809,0.841] | 0.790 [0.782,0.807] | 0.805 [0.794,0.820] |
| TRIAD | 124 | 0.627 [0.613,0.654] | 0.634 [0.612,0.662] | 0.634 [0.616,0.651] | 0.668 [0.649,0.702] | 0.590 [0.565,0.616] | 0.661 [0.640,0.679] | 0.632 [0.617,0.657] |
| WRAP | 82 | 0.636 [0.622,0.655] | 0.595 [0.541,0.620] | 0.904 [0.879,0.927] | 0.885 [0.861,0.921] | 0.843 [0.808,0.877] | 0.923 [0.902,0.948] | 0.904 [0.875,0.930] |

**Extended Figure 6.** Two-step approach for clinical trials using mPACC5 decline, with Tau-PET$_{NEO}$

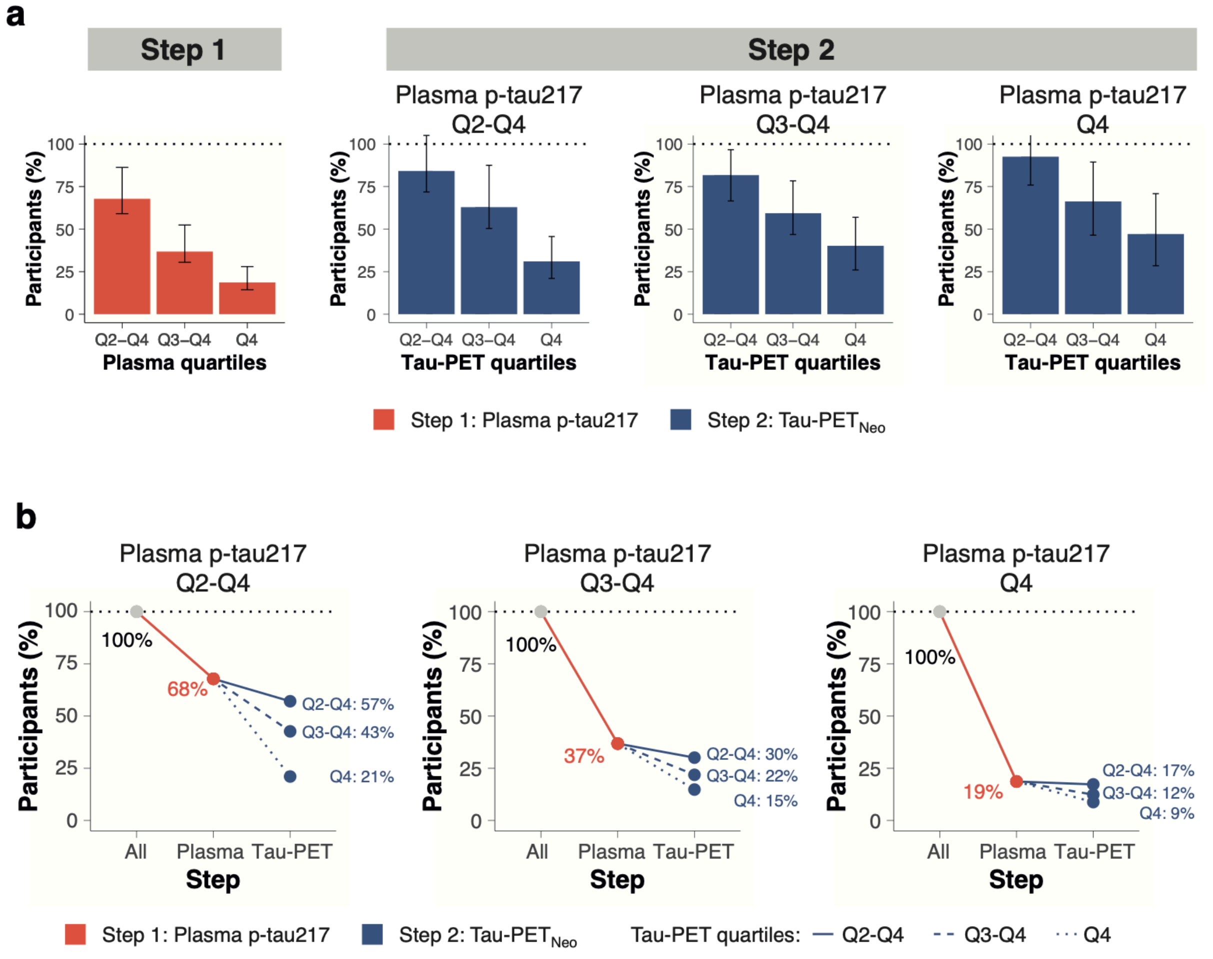


**a**, the obtained sample size reduction using different percentiles (75th, 50th and 25th) of the samples' baseline plasma p-tau217 baseline levels using the mPACC5 as the primary endpoint (step 1). Then, we repeated the approach selecting the 75th, 50th and 25th percentiles of the new samples' Tau-PET$_{NEO}$ measures (step 2). Note that 100% in step 2 refers to the participants selected by plasma p-tau217 in step 1. **b** shows the calculated sample size reductions for various plasma p-tau217 and Tau-PET$_{NEO}$ quantile combinations.

**Extended Figure 7.** Two-step approach for clinical trials using progression to MCI, with Tau-PET$_{NEO}$

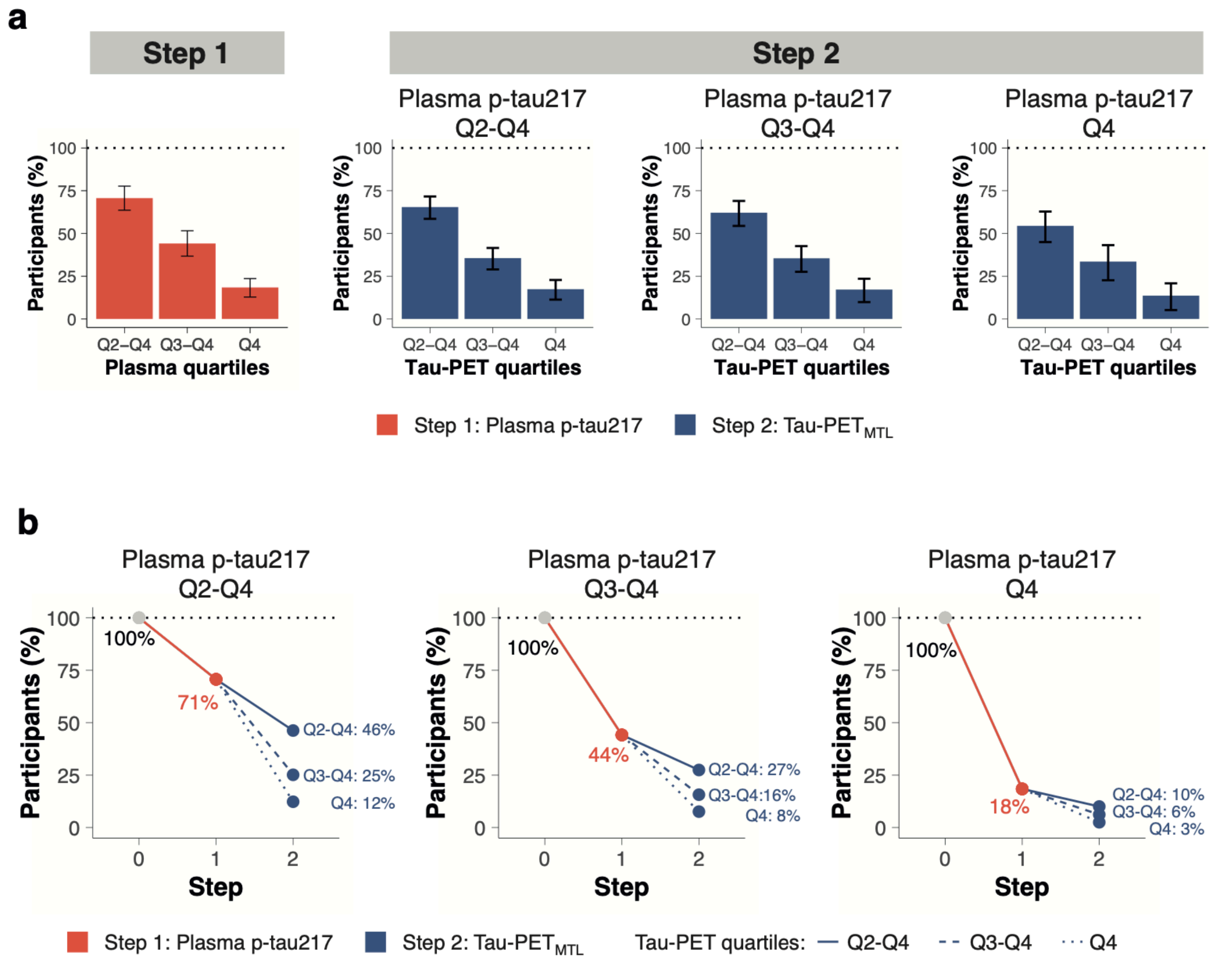


**a**, the obtained sample size reduction using different percentiles (75th, 50th and 25th) of the samples' baseline plasma p-tau217 baseline levels using progression to mild cognitive impairment as the primary endpoint (step 1). Then, we repeated the approach selecting the 75th, 50th and 25th percentiles of the new samples' Tau-PET$_{NEO}$ measures (step 2). Note that 100% in step 2 refers to the participants selected by plasma p-tau217 in step 1. **b** shows the calculated sample size reductions for various plasma p-tau217 and Tau-PET$_{NEO}$ quantile combinations.

**Extended Table 9.** Sample size reductions in a clinical trial following a two-step approach

| Step 1. Quantile Plasma | Step 2. Quantile PET | Plasma (%) | Tau-PET$_{MTL}$ (%) | Tau-PET$_{NEO}$ (%) | Tau-PET$_{MTL}$ (%, ref plasma) | Tau-PET$_{NEO}$ (%, ref plasma) |
|---|---|---|---|---|---|---|
| **Modified Preclinical Alzheimer Cognitive Composite 5 (mPACC5)** | | | | | | |
| **Q2-Q4** | Q2-Q4 | | 50[42, 70] | 57[49, 79] | 74[62, 94] | 84[72, 105] |
| | Q3-Q4 | | 35[31, 53] | 43[35, 64] | 52[44, 73] | 63[50, 88] |
| | Q4 | 68[59, 86] | 17[14, 27] | 21[15, 33] | 25[19, 37] | 31[21, 46] |
| **Q3-Q4** | Q2-Q4 | | 27[22, 40] | 30[24, 44] | 74[59, 88] | 82[67, 97] |
| | Q3-Q4 | | 19[15, 28] | 22[18, 33] | 50[39, 65] | 59[47, 78] |
| | Q4 | 37[31, 52] | 11[9, 18] | 15[11, 23] | 31[22, 44] | 40[26, 57] |
| **Q4** | Q2-Q4 | | 15[12, 23] | 17[13, 27] | 81[63, 102] | 93[76, 110] |
| | Q3-Q4 | | 10[8, 17] | 12[9, 20] | 56[40, 76] | 66[46, 89] |
| | Q4 | 19[14, 28] | 8[6, 13] | 9[6, 15] | 41[30, 63] | 47[28, 71] |
| **Clinical progression to mild cognitive impairment (MCI)** | | | | | | |
| **Q2-Q4** | Q2-Q4 | | 59[48, 69] | 46[39, 53] | 83[71, 94] | 65[59, 72] |
| | Q3-Q4 | 71[64, 78] | 43[32, 54] | 25[20, 30] | 61[47, 75] | 36[29, 42] |
| | Q4 | | 25[14, 35] | 12[8, 16] | 36[21, 49] | 17[11, 23] |
| **Q3-Q4** | Q2-Q4 | | 38[28, 46] | 27[22, 33] | 85[71, 98] | 62[54, 69] |
| | Q3-Q4 | 44[37, 52] | 30[19, 39] | 16[11, 20] | 68[47, 85] | 35[28, 43] |
| | Q4 | | 12[6, 17] | 8[4, 11] | 27[14, 38] | 17[10, 24] |
| **Q4** | Q2-Q4 | | 16[10, 22] | 10[7, 13] | 89[69, 108] | 54[45, 63] |
| | Q3-Q4 | 18[13, 24] | 12[6, 18] | 6[3, 9] | 66[40, 90] | 34[23, 43] |
| | Q4 | | 4[1, 6] | 3[1, 4] | 21[7, 32] | 14[5, 21] |

**Extended Figure 8.** Two-step approach for trials using mPACC5 decline in Aβ+ CU

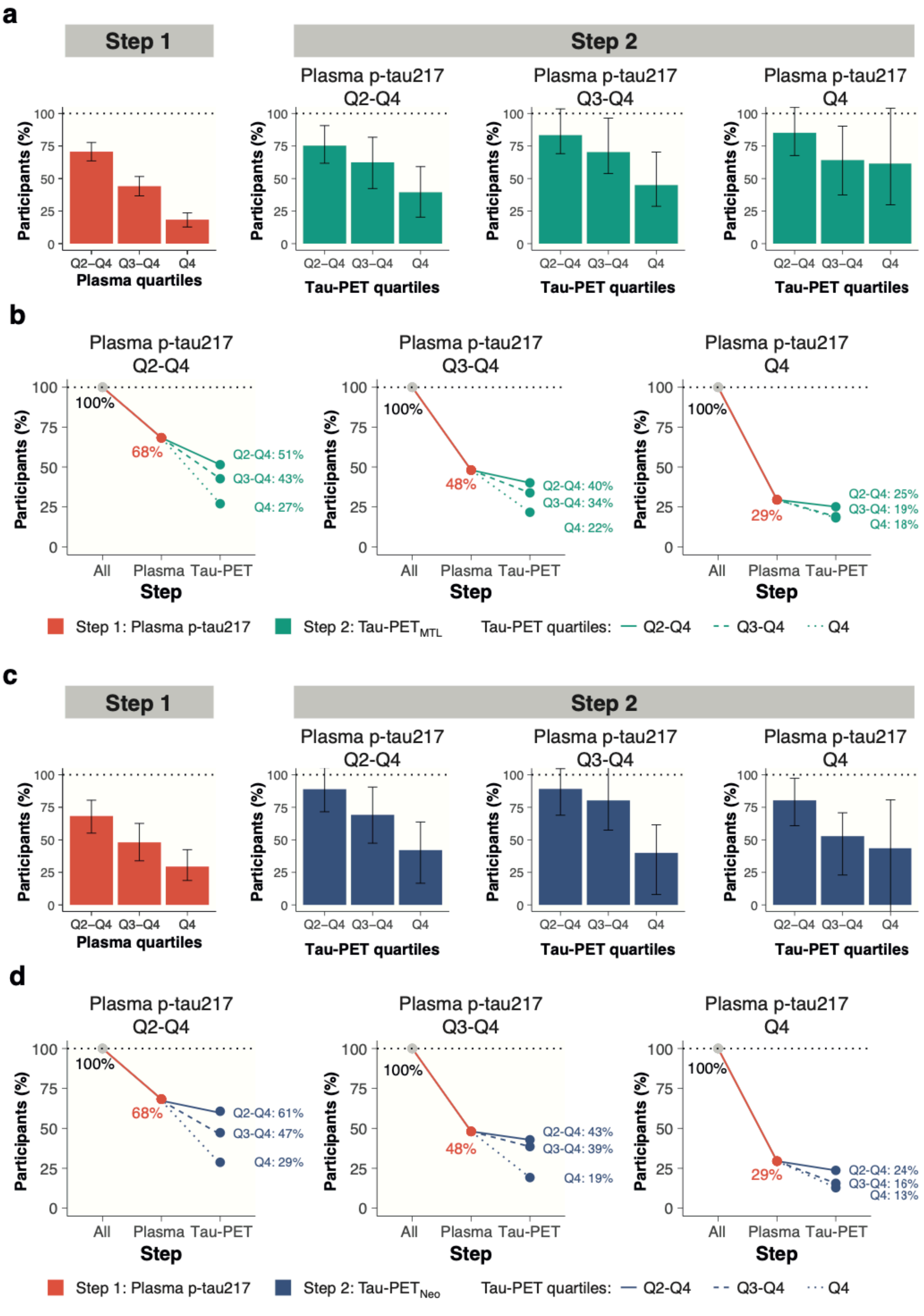


**a,c** the obtained sample size reduction using different percentiles (75th, 50th and 25th) of the samples' baseline plasma p-tau217 baseline levels using the mPACC5 as the primary endpoint (step 1). Then, we repeated the approach selecting the 75th, 50th and 25th percentiles of the new samples' Tau-PET$_{MTL}$ (**a**) or Tau-PET$_{NEO}$ (**c**) measures (step 2). Note that 100% in step 2 refers to the participants selected by plasma p-tau217 in step 1. **b,d** show the calculated sample size reductions for various plasma p-tau217 and samples' Tau-PET$_{MTL}$ (**b**) or Tau-PET$_{NEO}$ (**d**) quantile combinations.

**Extended Figure 9.** Characterization of different plasma p-tau217/Tau-PET$_{NEO}$ groups

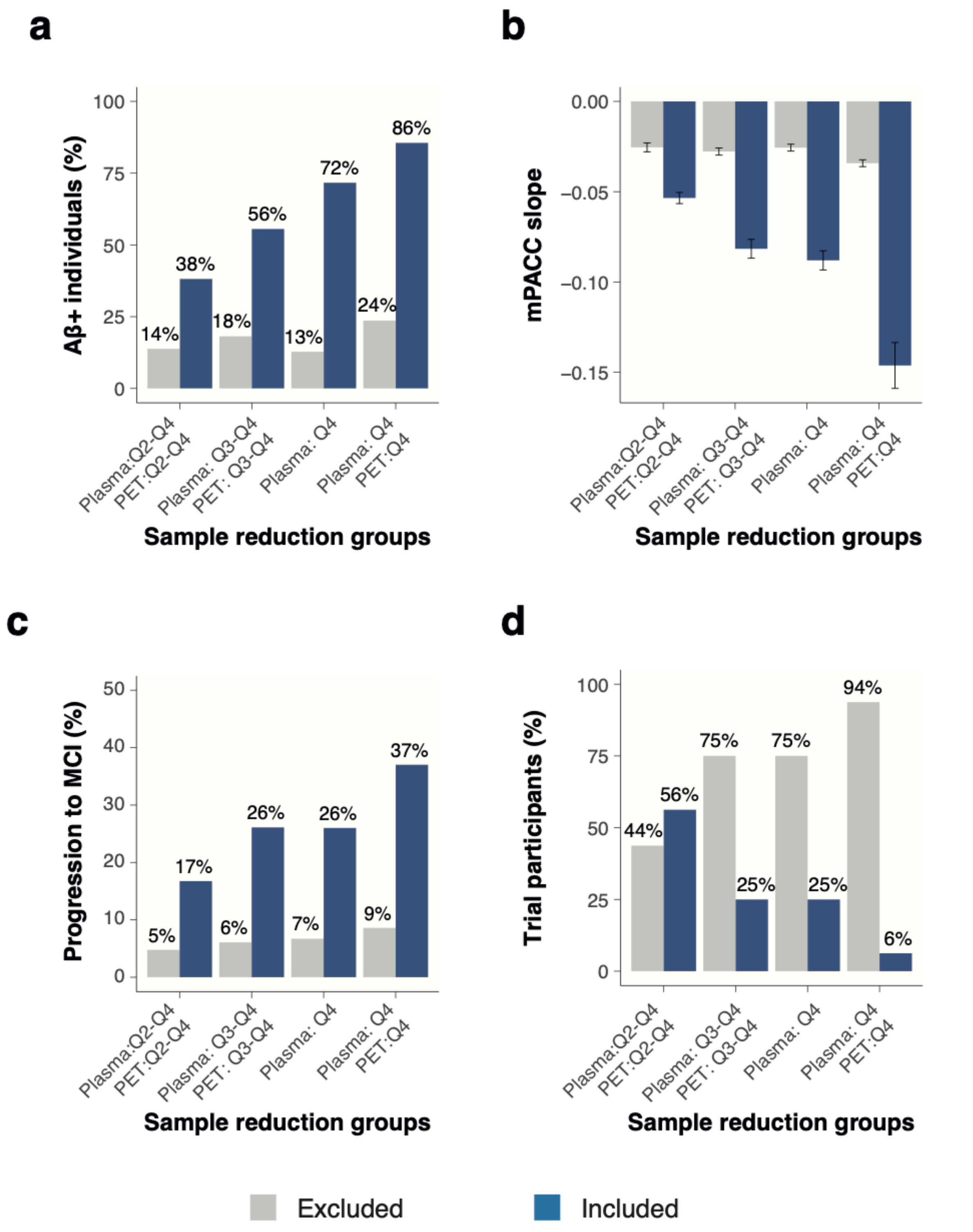


This figure shows how different group compositions based on their baseline plasma p-tau217 and Tau-PET$_{NEO}$ levels are related to various relevant trial metrics, including the proportion of Aβ+ individuals (**a**), annual mPACC5 slope (**b**), proportion of initially cognitively unimpaired individuals that progress to mild cognitive impairment during a 4-year trial (**c**), and the proportion of individuals from the entire population that fall within the group definitions described on the x-axis (**d**). Errorbars in **b** represent the 95% CI.

**Extended Table 10.** Combined plasma p-tau217 and Tau-PET$_{MTL}$ group characterizations: Aβ status and clinical outcomes

| | | | | INCLUDED POPULATION | | | EXCLUDED POPULATION | | |
|---|---|---|---|---|---|---|---|---|---|
| Plasma | PET | Excluded | Included | Aβ+ | mPACC slope | Progression to MCI | Aβ+ | mPACC slope | Progression to MCI |
| Q2-Q4 | All | 360 | 1080 | 34.1% | -0.05 (0.08) | 14.1% | 7.8% | -0.02 (0.06) | 3.8% |
| Q2-Q4 | Q2-Q4 | 630 | 810 | 39.8% | -0.06 (0.09) | 17.0% | 11.7% | -0.02 (0.06) | 4.1% |
| Q2-Q4 | Q3-Q4 | 900 | 540 | 47.2% | -0.07 (0.09) | 21.3% | 15.7% | -0.02 (0.06) | 5.2% |
| Q2-Q4 | Q4 | 1170 | 270 | 67.4% | -0.10 (0.10) | 29.5% | 18.3% | -0.03 (0.06) | 6.4% |
| Q3-Q4 | All | 720 | 720 | 46.5% | -0.06 (0.09) | 18.6% | 8.5% | -0.02 (0.06) | 4.4% |
| Q3-Q4 | Q2-Q4 | 900 | 540 | 53.0% | -0.07 (0.09) | 21.9% | 12.2% | -0.02 (0.06) | 4.6% |
| Q3-Q4 | Q3-Q4 | 1080 | 360 | 61.4% | -0.09 (0.10) | 26.5% | 16.2% | -0.03 (0.06) | 5.3% |
| Q3-Q4 | Q4 | 1260 | 180 | 81.1% | -0.12 (0.11) | 35.1% | 19.8% | -0.03 (0.07) | 6.4% |
| Q4 | All | 1080 | 360 | 71.7% | -0.09 (0.10) | 26.0% | 12.8% | -0.03 (0.06) | 6.7% |
| Q4 | Q2-Q4 | 1170 | 270 | 78.5% | -0.10 (0.10) | 29.0% | 15.7% | -0.03 (0.06) | 6.8% |
| Q4 | Q3-Q4 | 1260 | 180 | 87.2% | -0.12 (0.11) | 32.2% | 19.0% | -0.03 (0.07) | 7.2% |
| Q4 | Q4 | 1350 | 90 | 95.6% | -0.15 (0.12) | 40.1% | 23.0% | -0.03 (0.07) | 7.4% |

**Extended Table 11.** Combined plasma p-tau217 and Tau-PET$_{MTL}$ group characterizations: Demographic information

| | | | INCLUDED POPULATION | | | | | EXCLUDED POPULATION | | | |
|---|---|---|---|---|---|---|---|---|---|---|---|
| Plasma | PET | | Age | Females | Education | *APOE* ε4+ | | Age | % female | Education | *APOE* ε4+ |
| Q2-Q4 | All | | 70.5 (10.3) | 52.5% | 14.0 (3.3) | 40.5% | | 67.4 (10.2) | 57.2% | 14.2 (3.4) | 26.1% |
| Q2-Q4 | Q2-Q4 | | 71.7 (9.9) | 50.6% | 14.1 (3.4) | 41.7% | | 67.1 (10.4) | 57.6% | 14.0 (3.3) | 30.6% |
| Q2-Q4 | Q3-Q4 | | 73.4 (9.1) | 51.3% | 13.9 (3.3) | 44.8% | | 67.5 (10.4) | 55.1% | 14.1 (3.4) | 32.1% |
| Q2-Q4 | Q4 | | 74.9 (8.0) | 52.2% | 13.7 (3.4) | 49.3% | | 68.5 (10.4) | 54.0% | 14.1 (3.3) | 34.0% |
| Q3-Q4 | All | | 71.5 (10.4) | 50.6% | 14.0 (3.4) | 46.0% | | 67.8 (10.0) | 56.8% | 14.1 (3.3) | 27.8% |
| Q3-Q4 | Q2-Q4 | | 72.9 (9.8) | 49.8% | 13.9 (3.5) | 47.8% | | 67.7 (10.2) | 56.0% | 14.1 (3.3) | 30.3% |
| Q3-Q4 | Q3-Q4 | | 74.3 (9.2) | 50.3% | 13.8 (3.4) | 50.8% | | 68.1 (10.2) | 54.8% | 14.1 (3.3) | 32.2% |
| Q3-Q4 | Q4 | | 75.7 (7.7) | 55.6% | 13.6 (3.5) | 57.8% | | 68.8 (10.4) | 53.4% | 14.1 (3.3) | 33.9% |
| Q4 | All | | 73.8 (9.5) | 52.2% | 13.8 (3.5) | 51.4% | | 68.3 (10.3) | 54.2% | 14.1 (3.3) | 32.0% |
| Q4 | Q2-Q4 | | 75.1 (8.9) | 51.1% | 13.8 (3.5) | 54.4% | | 68.4 (10.2) | 54.3% | 14.1 (3.3) | 32.8% |
| Q4 | Q3-Q4 | | 75.6 (8.1) | 52.8% | 13.5 (3.5) | 59.4% | | 68.8 (10.4) | 53.8% | 14.1 (3.3) | 33.7% |
| Q4 | Q4 | | 74.1 (7.9) | 58.9% | 13.4 (3.4) | 63.3% | | 69.4 (10.4) | 53.3% | 14.1 (3.3) | 35.1% |

**Extended Figure 10.** Projected costs that could be saved in a hypothetical trial with mPACC5 as an endpoint

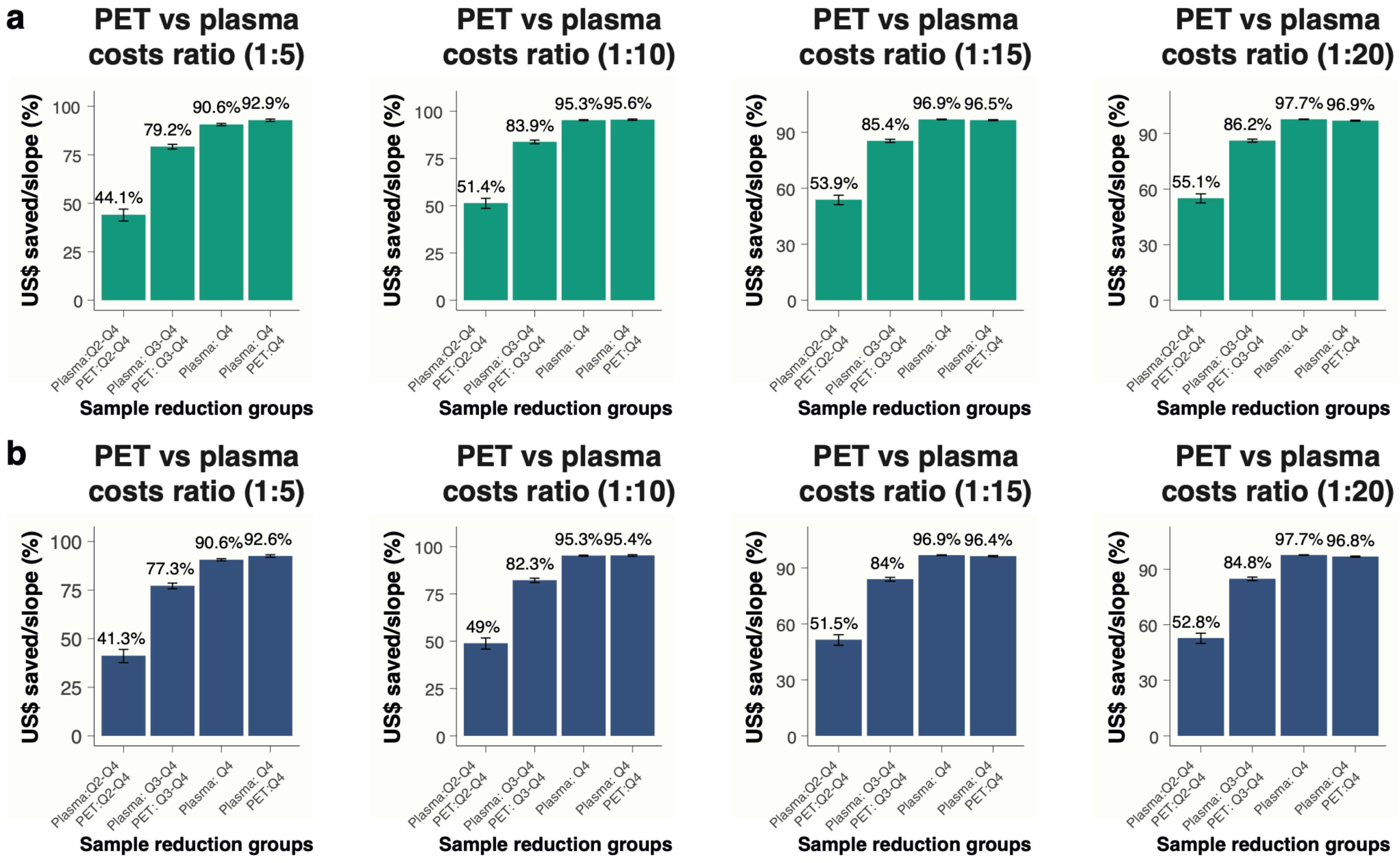


Figure shows the % of cost reductions that can be achieved when implementing different Tau-PET (Tau-PET$_{MTL}$ in panel **a,** Tau-PET$_{NEO}$ in panel **b**) vs plasma p-tau217 combinations when using the mPACC as an endpoint. The ratio of 1:5 reflects that the cost of 1 Tau-PET scan resembles the cost of 5 plasma p-tau217 assessment.

**Extended Figure 11.** Projected costs that could be saved in a hypothetical trial with clinical progression to MCI as an endpoint

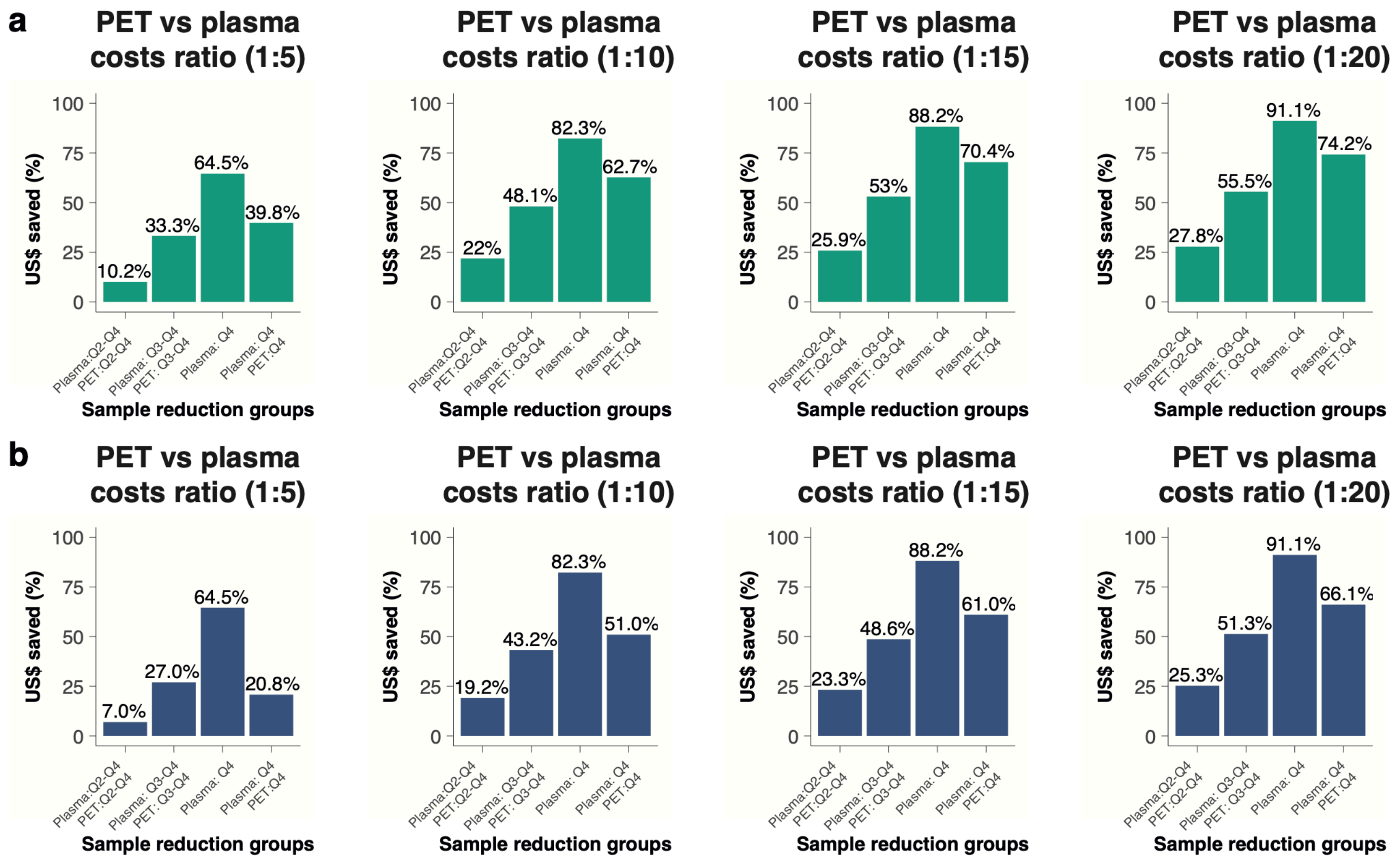


The % of cost reductions that can be achieved when implementing different Tau-PET (Tau-PET$_{MTL}$ in panel **a,** Tau-PET$_{NEO}$ in panel **b**) vs plasma p-tau217 combinations when using clinical progression to MCI as an endpoint. The ratio of 1:5 reflects that the cost of 1 Tau-PET scan resembles the cost of 5 plasma p-tau217 assessment.

**Extended Data Table 12.** Cohort descriptions

| Cohort | Cohort description | References |
|---|---|---|
| BioFINDER-1 & BioFINDER-2 | The Swedish BioFINDER studies are longitudinal studies covering the entire AD continuum in which participants were recruited at Skåne University Hospital and the Hospital of Angelholm, Sweden. The main inclusion criteria were absence of cognitive symptoms as assessed by a physician with special interest in cognitive disorders, being fluent in Swedish, having no significant unstable systemic illness that made it difficult to participate in the study, having no current significant alcohol or substance misuse, and no significant neurological or psychiatric illness. For the current study participants above > 50 years old were included. Both cognitively healthy older adults and SCD participants were included. The SCD participants were referred from participating memory clinic because of cognitive complaints, but did not fulfill criteria for MCI (defined using criteria by Petersen and operationalized according to[1,2]) following a neuropsychological test battery. | [3,4] |
| MCSA | The Mayo Clinic Study of Aging (MCSA) is a longitudinal population-based study of cognitive aging in Olmsted County, Minnesota. The study was designed to study prevalence, incidence and risk factors for MCI and dementia. Potential participants are randlomly enumerated from the Olmsted County, MN, census and enrolled by age/sex strata. Enumeration is repeated to maintain a sample of approximately 3000 active participants. At entry, every person underwent evaluations that included a medical history review and interview with the participant and a study partner, a neurological examination by a physician; and a neuropsychological examination. For this study, participants were considered MCI only if the study coordinator, physician, and neuropsychologist were all in agreement regarding the MCI diagnosis. Participants were judged cognitively normal if they did not meet MCI criteria. Participants aged between 50 and 89 years old were included in the current study. | [5] |
| Knight ADRC | The Charles F. and Joanne Knight Alzheimer Disease Research Center (Knight ADRC) is one of approximately 30 Centers funded by the National Institute on Aging (NIA) located at major medical institutions across the United States. Researchers at these Centers are working to translate research advances into improved diagnosis and care for people with Alzheimer disease, as well as working to find a treatment or way to prevent Alzheimer disease and other types of dementia. | [6] |
| PREVENT-AD | The PREVENT-AD (Pre-symptomatic Evaluation of Experimental or Novel Treatments for Alzheimer Disease) cohort is composed of cognitively healthy participants over 55 years old, at risk of developing Alzheimer Disease (AD) as their parents and/or siblings were/are affected by the disease. These | [7] |

| | | |
|---|---|---|
| | ‘at-risk’ participants have been followed for a naturalistic study of the presymptomatic phase of AD since 2011 using multimodal measurements of various disease indicators. Two clinical trials intended to test pharmaco-preventive agents have also been conducted. | |
| AIBL | The Australian Imaging, Biomarker & Lifestyle Flagship Study of Ageing (AIBL) is a longitudinal, prospective cohort with participants coming from two-site study – Melbourne and Perth. To be included in the study, participants were (1) ≥60 years old; (2) fluent in English; (4) had completed at least 7 years of education; (5) did not have any history of neurological or psychiatric disorders, drug or alcohol abuse or dependence, or any other unstable medical condition; and (6) were deemed to be cognitively unimpaired (CU), based on their performance on a battery of cognitive assessments that AIBL participants undergo every 12 to 18 months. A multidisciplinary clinical review panel determines whether an individual is CU, based on the available clinical and neuropsychological information. | 8 |
| ADC | The Amsterdam Dementia Cohort (ADC) is a prospective cohort study including (amongst others) individuals with subjective cognitive decline (SCD) presenting at the Alzheimer Center of the VU University Medical Center Amsterdam. All participants have been referred to the memory clinic by their general practitioner, and a neurologist or geriatrician in the case of a second opinion for evaluation of cognitive complaints. They receive standardized dementia screening at the memory clinic, including an interview with a neurologist, physical and neurological examination, neuropsychological assessment. Individuals with SCD can additionally be included in the SCIENCe study, for which the main inclusion criteria are a diagnosis of SCD (i.e., cognitive complaints and normal cognition) and age ≥ 45 years. Exclusion criteria for participation in the SCIENCe study are MCI, dementia, major psychiatric disorder (i.e., current depression, personality disorders, schizophrenia), neurological diseases known to cause memory complaints (i.e., Parkinson’s disease, epilepsy), HIV, abuse of alcohol or other substances, and language barrier. | 9 |
| WRAP | The Wisconsin Registry for Alzheimer's Prevention is a longitudinal observational cohort study enriched with persons with a parental history (PH) of probable Alzheimer's disease (AD) dementia. Recruitment sources included memory clinics in which a parent was diagnosed or treated, limited radio and newspaper advertisements, and word of mouth. Participants generally meet the following inclusion criteria at study entry: age 40–65 years; fluent English speaker; visual and auditory acuity adequate for neuropsychological testing; good health with no diseases expected to interfere with study participation over time. Participants are excluded from enrollment if they have a prior diagnosis of dementia or evidence of dementia at baseline testing (one was excluded due to baseline dementia). | 10 |

| | | |
|---|---|---|
| TRIAD | The Translational Biomarkers of Aging and Dementia (TRIAD) cohort study is a longitudinal observational cohort study in Montréal, Québec, Canada. Participants are recruited from the community and from the the McGill Centre for Studies in Aging. All participants are clinically evaluated by dementia specialists. Participants were excluded from this study if they had systemic conditions which were not adequately controlled through a stable medication regimen. Other exclusion criteria were active substance abuse, recent head trauma, recent major surgery, or MRI/PET safety contraindications. The study was approved by the Montreal Neurological Institute PET working committee and the Douglas Mental Health University Institute Research Ethics Board. Written informed consent was obtained for all participants. | [11] |

**Extended Data Table 13.** Methods to determine Amyloid PET status by cohort

| Cohort | Tracer | Methodology | Cut-off | References |
|---|---|---|---|---|
| BioFINDER-1 | [$^{18}$F]flutemetamol | Global neocortical composite standardized uptake value ratios (SUVR) for the 90-110min interval p.i. with whole cerebellum as reference region | >1.03 SUVR | [4] |
| BioFINDER-2 | [$^{18}$F]flutemetamol | Global neocortical composite SUVR for the 90-110min interval p.i. with whole cerebellum as reference region | >1.03 SUVR | [4] |
| MCSA | [$^{11}$C]PIB | Late uptake amyloid PET images were acquired from 40-60 minutes p.i. A meta-ROI was calculated as the voxel-number weighted average of uptake in a target region including prefrontal, orbitofrontal, parietal, temporal, anterior and posterior cingulate, and precuneus regions divided by the uptake in the cerebellar crus gray matter. | >1.48 SUVR (>21CL) | [5] |
| Knight ADRC | [$^{11}$C]PIB | Data were processed using a region of interest approach using Freesurfer. Amyloid deposition was summarized using the average across the left and right lateral orbitofrontal, medial orbitofrontal, rostral middle frontal, superior frontal, superior temporal, middle temporal, and precuneus regions. | >20 CL | [6] |
| PREVENT-AD | [$^{18}$F]NAV4694 | Aβ-PET images were realigned onto their respective MRI, masked to remove the scalp and CSF in an attempt to avoid contamination by nongray or nonwhite matter voxels, and smoothed using a full width at half maximum Gaussian kernel of 8mm. Resulting images were scaled using whole cerebellum uptake values (whole cerebellum was preferred to cerebellum gray matter to account better for white matter off-target binding variability between tracers). Global neocortical Aβ burden was quantified by extracting, in native space, the mean standardized uptake value ratio (SUVR) of the frontal, temporal, parietal, and posterior cingulate cortex of the Desikan-Killiany atlas | >1.33 SUVR | [12] |
| AIBL | [[$^{18}$F]NAV4694 | The standard Centiloid (CL) cortical and whole cerebellar volumes of interest template were applied to the summed and spatially normalised PET images in order to obtain SUVR's. These SUVR were transformed into CL units by linear transformation using the PET tracer-specific equations published for conversion of CL method SUVR to CL units. | >24 CL | [13] |

| | | | | |
|---|---|---|---|---|
| ADC | [$^{18}$F]florbetapir | Visual read following guidelines provided by Avid Radiopharmaceuticals corresponding to >17 CL. | - | [14] |
| WRAP | [$^{11}$C]PIB | Amyloid burden was assessed as a global average $^{11}$C-PiB distribution volume ratio (DVR; Logan graphical analysis, cerebellum gray matter reference region), taken across 8 bilateral cortical ROIs. A+ was ascertained using a global $^{11}$C-PiB DVR≥1.16 a threshold previously shown to predict subsequent amyloid accumulation. | >1.16 DVR | [15] |
| TRIAD | [$^{18}$F]NAV4694 | [$^{18}$F]AZD4694 PET images were acquired 40-70 min after bolus injection and reconstructed on a 4-dimensional volume with 3 frames (3 x 600s). Amyloid-β SUVR from a neocortical region of interest (ROI) for each participant was estimated by averaging the SUVR from the precuneus, prefrontal, orbitofrontal, parietal, temporal, and cingulate cortices, with amyloid-β positivity defined as an [$^{18}$F]AZD4694 above 1.55. | >1.55 SUVR | [16] |

CL = Centiloid; DVR = Distribution volume ratio; SUVR = Standardized uptake value ratio.

Centiloid (CL) units were presented when available.

**Extended Data Table 14.** Methods to determine Tau PET status in the medial temporal lobe (MTL) and neocortex (NEO) by cohort

| Cohort | Tracer | Scanning interval | Reference region | Reference |
|---|---|---|---|---|
| BioFINDER-1 | [$^{18}$F]flortaucipir | 80-100min p.i. | Inferior cerebellar GM | [17] |
| BioFINDER-2 | [$^{18}$F]RO948 | 70-90min p.i. | Inferior cerebellar GM | [18] |
| MCSA | [$^{18}$F]flortaucipir | 80-100min p.i. | Cerebellar crus GM | [19] |
| Knight ADRC | [$^{18}$F]flortaucipir | 80-100min p.i. | Cerebellar GM | [6] |
| PREVENT-AD | [$^{18}$F]flortaucipir | 80-100min p.i. | Inferior cerebellar GM | [7] |
| AIBL | [$^{18}$F]MK6204 | 90-110 min p.i. | Cerebellar GM | [13] |
| ADC | [$^{18}$F]flortaucipir | 80-100min p.i. | Cerebellar GM | [20] |
| WRAP | [$^{18}$F]MK6240 | 70-90min p.i. | Inferior cerebellar GM | [15] |
| TRIAD | [$^{18}$F]MK6240 | 90-100min p.i. | Cerebellar Crus GM | [21] |

GM = Gray matter; MTL = Medial temporal lobe; NEO = Neocortical; p.i. = Post-injection; SUVR = Standardized uptake value ratio.

The cut-offs were generated in each individual cohort, based on the mean + 2*standard deviation across all Aβ-negative participants within each cohort. We computed tau PET status for a medial temporal lobe (MTL; unweighted average of bilateral entorhinal cortex and amygdala) and a neocortical (NEO; weighted average of bilateral middle temporal and inferior temporal gyri) region-of-interest.

**Extended Data Table 15.** Composition of the mPACC5 for each cohort

| Cohort | Global Cognition | Episodic Memory | Time executive function | Semantic memory |
|---|---|---|---|---|
| BioFINDER-1 | MMSE | ADAS-COG delayed word recall | Symbol digit modalities test | Animal fluency |
| BioFINDER-2 | MMSE | ADAS-COG delayed word recall | Symbol digit modalities test | Animal fluency |
| MCSA | MMSE[a] | AVLT delayed recall | WAIS-R Digit Symbol | Sum of animal, fruits and vegetables fluency |
| Knight ADRC | MMSE | CVLT – Delayed recall | Symbol digit modalities test | Animal fluency |
| PREVENT-AD | MMSE | SRT – Delayed recall | Symbol digit modalities test | Animal fluency |
| AIBL | MMSE | CVLT – Delayed recall | Symbol digit modalities test | Sum of animal and names fluency |
| ADC | MMSE | RAVLT – Delayed recall | TMT-B | Animal fluency |
| WRAP | MMSE | AVLT – Delayed recall | WAIS-R Digit Symbol | Animal fluency |
| TRIAD | MMSE | Logical Memory test - Delayed recall | Letter fluency | Category fluency |

Note that the episodic memory test was given double weight and thus accounted for 40% of the mPACC5 score.

[a] A 38-point test, the Short Test of Mental Status (STMS)[22], was converted to MMSE scores using an in-house developed algorithm[23].
